# *home*RNA self-blood collection enables high-frequency temporal profiling of presymptomatic host immune kinetics to respiratory viral infection: a prospective cohort study

**DOI:** 10.1101/2023.10.12.23296835

**Authors:** Fang Yun Lim, Hannah G. Lea, Ashley Dostie, Soo-Young Kim, Tammi van Neel, Grant Hassan, Meg G. Takezawa, Lea M. Starita, Karen Adams, Michael Boeckh, Joshua T. Schiffer, Ollivier Hyrien, Alpana Waghmare, Erwin Berthier, Ashleigh B. Theberge

**Author notes:** Co-senior authors: Ashleigh B. Theberge, Erwin Berthier, Alpana Waghmare.

## Abstract

**Background:** Early host immunity to acute respiratory infections (ARIs) is heterogenous, dynamic, and critical to an individual’s infection outcome. Due to limitations in sampling frequency/timepoints, kinetics of early immune dynamics in natural human infections remain poorly understood. In this nationwide prospective cohort study, we leveraged a Tasso-SST based self-blood collection and stabilization tool (*home*RNA) to profile detailed kinetics of the presymptomatic to convalescence host immunity to contemporaneous respiratory pathogens.

**Methods:** We enrolled non-symptomatic adults with recent exposure to ARIs who subsequently tested negative (exposed-uninfected) or positive for respiratory pathogens. Participants self-collected blood and nasal swabs daily for seven consecutive days followed by weekly blood collection for up to seven additional weeks. Symptom burden was assessed during each collection. Nasal swabs were tested for SARS-CoV-2 and common respiratory pathogens. 92 longitudinal blood samples spanning the presymptomatic to convalescence phase of eight SARS-CoV-2-infected participants and 40 interval-matched samples from four exposed-uninfected participants were subjected to high-frequency longitudinal profiling of 785 immune genes. Generalized additive mixed models (GAMM) were used to identify temporally dynamic genes from the longitudinal samples and linear mixed models (LMM) were used to identify baseline differences between exposed-infected (n = 8), exposed-uninfected (n = 4), and uninfected (n = 13) participant groups.

**Findings:** Between June 2021 – April 2022, 68 participants across 26 U.S. states completed the study and self-collected a total of 691 and 466 longitudinal blood and nasal swab samples along with 688 symptom surveys. SARS-CoV-2 was detected in 17 out of 22 individuals with study-confirmed respiratory infection, of which five were still presymptomatic or pre-shedding, enabling us to profile detailed expression kinetics of the earliest blood transcriptional response to contemporaneous variants of concern. 51% of the genes assessed were found to be temporally dynamic during COVID-19 infection. During the pre-shedding phase, a robust but transient response consisting of genes involved in cell migration, stress response, and T cell activation were observed. This is followed by a rapid induction of many interferon-stimulated genes (ISGs), concurrent to onset of viral shedding and increase in nasal viral load and symptom burden. Finally, elevated baseline expression of antimicrobial peptides were observed in exposed-uninfected individuals.

**Interpretation:** We demonstrated that unsupervised self-collection and stabilization of capillary blood can be applied to natural infection studies to characterize detailed early host immune kinetics at a temporal resolution comparable to that of human challenge studies. The remote (decentralized) study framework enables conduct of large-scale population-wide longitudinal mechanistic studies.

**Funding:** This study was funded by R35GM128648 to ABT for in-lab developments of *home*RNA and data analysis, a Packard Fellowship for Science and Engineering from the David and Lucile Packard Foundation to ABT, and R01AI153087 to AW.

## INTRODUCTION

Host innate and acquired immunity is critical to the outcomes of ARIs^1^. Systemic dysregulation of pro-inflammatory molecules is associated with ongoing inflammation and poor outcomes, while effective coordination of early responses limits the severity and duration of symptoms^2^. The dynamics of early host responses soon after pathogen exposure coupled with pre-existing host immunity at the time of exposure can pivot the host into either abortive or productive infection, and early viral clearance or progression to prolonged symptomatic disease. However, detailed kinetics of early immune responses especially during the presymptomatic period are poorly understood, including those conferring protective immunity in exposed individuals who remain asymptomatic or virology negative (exposed-uninfected). As in-person venipuncture presents a significant logistical barrier to longitudinal studies requiring early and frequent samples, prior studies of natural ARIs were largely cross-sectional, were primarily focused on post-symptomatic infections, typically when nasal viral loads were already in decline, and were often limited to infrequent sampling across inconsistent collection intervals^3-6^.

High-frequency longitudinal profiling of host peripheral immunity, particularly during extremely early presymptomatic phases, has largely been conducted in the setting of human challenge studies^7-9^. Controlled human infection models pose limitations to understanding the host response during natural infections^10,11^. Restricted by ethical and regulatory considerations, these models often employ use of a limited set of challenge strains, a uniform viral challenge dose, and are performed on low-risk, healthy volunteers, thus limiting their broad applicability to contemporaneously circulating variants and to the heterogeneity of population-wide immunologic backgrounds that are shaped by individual susceptibilities, and by prior vaccination and infection^11^. Studies of the host blood transcriptional response during presymptomatic phase of natural infections are very limited^12-14^ — with the first of such studies published by McClain *et al*. in 2021 that measured the presymptomatic host response to pre-COVID-19 respiratory viruses through daily monitoring of exposed close contacts residing in college dormitories^12^. During the COVID-19 pandemic, Gupta *et al*, in 2021 measured the presymptomatic host response to SARS-CoV-2 through weekly surveillance of healthcare workers^13^. In the first SARS-CoV-2 human challenge study using a pre-alpha challenge strain of SARS-CoV-2 published in March 2022, detectable nasal viral load (viral shedding) occurred between 40-76 hours post challenge followed shortly by onset of symptoms, making the presymptomatic phase of SARS-CoV-2 infection from natural exposures particularly hard to capture and characterize^15^. Thus, high-frequency daily measurements of the early host response to natural SARS-CoV-2 infection particularly in the latter variants of concern are unprecedented prior to this study.

A study framework that enables frequent longitudinal measurements, in a natural field setting, of the early host immune response to respiratory infections acquired through community transmissions can be integrated into the study design of future human challenge trials involving natural exposures to pathogens. Challenge by natural exposure to infected persons has recently been proposed as future designs of human challenge trials for vaccine development to circumvent dose-escalation studies, and address limitations surrounding strain selection, diversity, and delays in manufacturing timelines^16^. The ability to rapidly disseminate self-collection kits to recently exposed individuals across a wide geographic area (decentralized sampling) coupled with frequent self-collections at pre-scheduled infection timepoints and immediate on-site stabilization of the transcriptome is integral to conducting population-wide longitudinal mechanistic studies aimed at profiling detailed host immune kinetics during early and highly dynamic stages of natural infections (**Figure 1**). Our team recently developed *home*RNA, a tool for home transcriptomic studies that integrates capillary blood collection (Tasso-SST) with immediate RNA stabilization using RNA*later* in a custom engineered stabilizer tube with integrated fluidics^17^. *home*RNA enables self-collection, on-site stabilization of liquid capillary blood (up to 0.5 mL volume), and ambient temperature mailing of stabilized liquid capillary blood back to a central lab^17,18^; we recently demonstrated its capability of capturing SARS-CoV-2 infection signatures in non-hospitalized adults and discriminating distinct infection response between previously vaccinated and unvaccinated individuals^19^. Here, we aim to assess the feasibility of using self-collection methodologies (*home*RNA and nasal swab) within a fully remote longitudinal study setting to profile, at deep temporal resolution, the presymptomatic immune kinetics to community-acquired ARIs in recently exposed individuals recruited across the United States. Despite the small cohort size as a feasibility study, to the best of our knowledge, this is the first prospective longitudinal nationwide study to assess the presymptomatic blood transcriptional response to a natural respiratory infection at a temporal resolution comparable to that of human challenge studies. The study design and framework presented here can be adapted to augment measurements of dynamic immune responses in future clinical studies.

**Figure 1.**
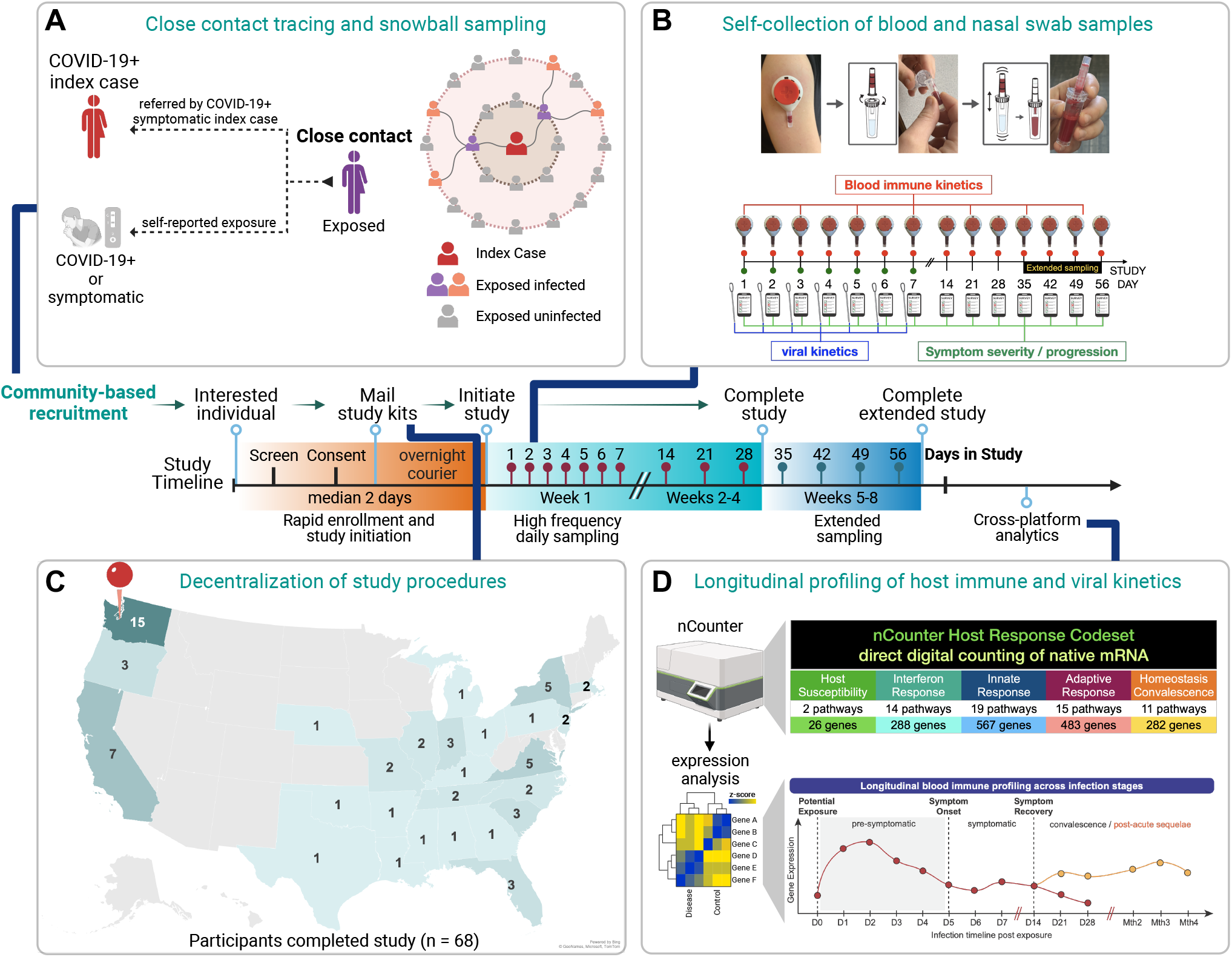
Study framework using home self-collection tools to observe detailed kinetics of early presymptomatic host transcriptional response during natural human respiratory infections. Conceptual framework depicting the study design and participation timeline (middle), A) close contact recruitment strategies, B) *home*RNA, nasal swab, and symptom survey collection details (Tasso-SST blood collection and blood RNA stabilization process adapted from Haack and Lim et al, 2021 with permission), C) geographical representation of study participants, and D) longitudinal gene expression analyses pipeline.

## METHODS

### Study design

This is a prospective longitudinal study aimed at assessing the feasibility of home self-collection methodologies to profile detailed kinetics of extremely early (presymptomatic) host immunity to respiratory pathogens with a focus on SARS-CoV-2 in non-hospitalized adults using self-blood (*home*RNA) and nasal swab collection methodologies. Between June 2021 April 2022, individuals who had a recent exposure to those with a confirmed (SARS-CoV-2 nucleic acid or antigen test) or suspected (symptomatic) COVID-19 infection and who were also asymptomatic at the time of screening were recruited from the general population throughout the United States. Each participant self-collected 10-14 blood and seven nasal swab samples over 4-8 weeks. Symptom burden was assessed on each collection day. The study was approved by the University of Washington (UW) IRB (STUDY00012546) and conducted at the UW. All participants provided informed consent. All samples were collected remotely by study participants and all surveys were administered through REDCap^20^.

### Participants

Participants were recruited via social media (Facebook) and news platforms (WATE-TV), the Institute of Translational Health Sciences Participate-in-Research program, and the UW Husky Coronavirus Testing Program sites. Multiple recruitment strategies were employed to enrich for infected individuals who were still within their presymptomatic phase of infection: adult close contacts with unknown infection status and asymptomatic at the time of screening were recruited directly via i) self-reported exposure or ii) referral by symptomatic SARS-CoV-2 positive individuals (index cases) (**Figure 1A**). Index cases were asked to refer their adult close contacts who gave permission. In addition, the study also enrolled COVID-19+ individuals with known self-reported SARS-CoV-2 infection status who were still asymptomatic at the time of screening. Screening surveys were administered online through REDCap. Eligible asymptomatic close contacts and COVID-19+ individuals who provided informed consent were enrolled into the study for serial sampling of both blood and nasal swab samples as described below. For this feasibility study, participants were enrolled in two waves (June - October 2021; January April 2022) until at least 15 participants with confirmed ARIs were reached. Longitudinal immune profiles of unexposed-healthy participants were obtained from a previously published study that utilized the same *home*RNA collection and analysis platform^19^. Specifically, 5 healthy volunteers collected longitudinal samples every other day for two weeks. In addition, 8 healthy volunteers collected single timepoint samples.

### Procedures

A study package containing *home*RNA blood (Tasso-SST blood collection device and RNA stabilizing tube)^17^ and nasal swab collection kits, prepaid return mailers, an informational packet with all instructions for use, a collection calendar with pre-scheduled collection dates, and specimen return instructions were mailed to study participants, who performed their first collection the day of package receipt. All participants collected blood and nasal swab samples daily for seven consecutive days (study days 1-7) followed by weekly blood collection for three additional weeks (study days 14, 21, 28) (**Figure 1B**). A subset of study-confirmed COVID-19+ participants were selected to extend blood collections for up to four additional weeks (study days 35, 42, 48, 56). On each collection date, participants were prompted with an email or text message containing a link to the online REDCap survey to fill out a device use and symptom burden survey. The first nasal swab from each participant was submitted for clinical SARS-CoV-2 RT-PCR, and test results were returned to participants who enrolled as close contacts with unknown infection status. 3-4 additional nasal swabs were selected based on symptom burden on the day of collection to screen for 18 common respiratory pathogens through a lab-developed Multi-pathogen OpenArray assay (**Table S1**). For participants with positive SARS-CoV-2 results in research swab samples, one representative sample with the lowest Crt value (highest viral load) was selected for SARS-CoV-2 sequencing for variant identification. Detailed immune kinetics of 773 innate and acquired immune genes across 57 pathways associated with the host response to infection and 12 housekeeping genes (nCounter® Host Response Panel) were profiled using nCounter® direct digital counting of native RNA (**Figure 1D**)^19^. Detailed methods are described in the supplemental methods. Participants were classified based on their self-reported COVID-19 infection status at enrollment and virology status of collected nasal swabs (**Figure S1A**): 1) participants positive for SARS-CoV-2 were classified as “COVID-19+”; 2) participants negative for SARS-CoV-2 but positive for other respiratory pathogens were classified as “other virus”; 3) participants negative for all respiratory pathogens were classified as “exposed-uninfected”;4) participants negative for SARS-CoV-2 through testing of research samples but self-reported a recent SARS-CoV-2 positive test were classified as “COVID-19 probable”. COVID-19 probable participants who initiated their first collection > 14 days after their most recent positive test date were classified as “COVID-19 convalescent”.

### Symptom burden and severity scoring

A modified Jackson score was used to compute the daily symptom burden ^8,12^. Specifically, participants were asked to rate the severity for each surveyed symptom based on assigned severity scores of 0-3 (0 = no symptom, 1= mild, 2 = moderate, and 3 = severe). The total sum daily score of 26 symptoms was used to compute daily symptom severity scores (range: 0-78). The day of symptom onset was defined by the first of two consecutive days with a total sum daily score of 2 and used to distinguish presymptomatic from early acute infections (**Figure S1B**). The average symptom severity score for the first seven days of sampling was used to define total symptom severity and distinguish symptomatic and asymptomatic infections. Participants with a total severity score of < 1 were defined as asymptomatic.

### Statistical analysis

Detailed methods on RNA isolation, gene expression analysis, normalization procedures, and functional enrichment analysis of differentially expressed transcripts are described in supplemental methods. All references for statistical analyses described below can be found in the supplemental methods. nCounter gene expression data available as normalized counts in **supplemental data 1**. See supplemental info for references used in statistical analyses.

Generalized additive mixed models (GAMM): Gene expression was modeled as trimmed mean of means (TMM) normalized log2 counts per million. Generalized additive mixed models (GAMM were used to evaluate associations between gene expression and disease status (healthy, exposed-uninfected, and COVID-19+) over time. These analyses accounted for potential confounding factors (age, sex, and nCounter Host Response codeset versions), integrated subject-specific random intercepts to accommodate for potential within-subject correlation, and integrated smooth functions of time using a sample’s collection date relative to the individual’s first PCR positivity date (s(day PVS)). Smooth functions obtained by the GAMM were used to characterize longitudinal patterns of gene expression over time, ranging from four days prior to the initial positive PCR test (D-4) to 29 days afterward (D29) for COVID-19+ subjects. For healthy individuals and those exposed but uninfected, the data were recorded between the day of the initial sampling (D1) and 29 days thereafter (D29). P-values were adjusted for multiple comparisons by controlling the false discovery rate (FDR) using the Benjamini-Hochberg (BH) procedure. GAMM analyses with thin plate regression splines with 6 basis dimensions were carried out using the gamm4 R package. Genes with adjusted p-values < 0.1 for the smooth term of time (s(day PVS)) in COVID-19+ participants were changing over time and considered COVID-19 dynamic genes. Genes with adjusted p-values < 0.05 were considered significant while genes with adjusted p-values between 0.05 – 0.1 were considered trending. GAMM analysis results are summarized in **supplemental data 2**.

Identification of time-trend clusters: Hierarchical clustering based on the Spearman’s rank correlation between gene-specific temporal profiles derived from the fitted GAMM was used to group genes exhibiting similar longitudinal patterns. A total of 324 significant COVID-19 dynamic genes with BH-adjusted p-values < 0.05 for the GAMM smooth term of time (s(day PVS)) for the COVID-19+ group were subjected to hierarchical clustering. Next, hierarchical clustering with complete linkage based on the correlation matrix of these 324 genes was performed to group genes sharing similar longitudinal trends. The analysis identified a total of eleven time-trend clusters (C1-C11; **supplemental data 3**).

Linear mixed models (LMM): We examined the influence of disease status on gene expression during the earliest stages of the disease using LMM within the kimma R package. Three disease status groups were assessed: unexposed-uninfected (healthy), exposed-uninfected (exposed), and exposed-infected (COVID-19+). The analyses adjusted for disease status, time (day PVS), and the interaction between disease status and time (day PVS), as well as potential confounding factors (age, sex, and nCounter Host Response codeset versions). Gene-level quality weights were calculated using voomWithQualityWeights in limma R package and incorporated into the model to account for gene-level variability between different observations. Subject-specific random intercepts and slopes for the variable “day PVS” were included in the model to accommodate potential within-subject correlation and variability in the relationship between gene expression and time. Longitudinal samples from the day of initial PCR positivity (day PVS = D0) to five days thereafter (day PVS = D5) were used. In healthy and exposed-uninfected individuals, these correspond to the day of the initial sampling (D1) up to five days thereafter (D5). To identify differentially expressed genes between disease groups, 261 genes with adjusted p values < 0.05 in the main disease variable were subjected to pairwise contrasts between two disease groups as follow: “exposed vs healthy”, “COVID-19+ vs healthy”, and “COVID-19+ vs exposed”. P-values were adjusted for multiple comparisons by controlling the FDR using the BH procedure. LMM analysis results are summarized in **supplemental data 4**.

Gene ontology overrepresentation analyses: 324 COVID-19 dynamic genes with BH-adjusted p-values < 0.05 were grouped into early pre-shedding (Temporal trends 1 and 2; N = 69), early post-shedding (Temporal trend 3; N = 158), and mid-to-late genes (Temporal trend 4; N = 97). Overrepresentation analyses were performed on both early pre-shedding and post-shedding genes using the enrichGO() function of the clusterProfiler R package. The human GO biological process database (org.Hs.eg.db) was used to identify pathways enriched in each group. GO terms consisting of >=5 genes from the initial query list and a BH-adjusted p-value of < 0.05 were subjected to parent term analysis to reduce term redundancy and facilitate biological interpretation of enriched pathways. Specifically, 201 and 425 significant GO terms associated with pre-and post-shedding genes respectively were used in parent term analysis. For parent term analyses, semantic similarity between pairs of GO terms were calculated and a matrix of dissimilarities was computed using the calculateSimMatrix() function of the rrvgo package. The terms were clustered using complete linkage and reduced to parent term clusters using the reduceSimMatrix() function at a threshold of 0.7.

Geneset enrichment analysis (GSEA): GSEA was performed using the gsePathway() function of the ReactomePA R Package. Comparison of gene expression between COVID-19+ and Exposed were performed on all 785 genes using the LMM analysis (see LMM methods section above). Resulting estimates of the average difference between gene expression measured in COVID-19+ and in Exposed (reference) were used to rank-order genes from highest (overexpressed in COVID-19+) to lowest (overexpressed in Exposed). P-values were adjusted for multiple comparisons using the BH procedure and an adjusted p-value < 0.1 was used to identify enriched pathways.

## RESULTS

Between June 2021 – April 2022, 90 non-symptomatic adults with recent exposure to ARIs were enrolled for high-frequency serial self-sampling of peripheral blood using *home*RNA and nasal swabs (**Figure 2**; see Table S1 for ineligibility reasons). Participants self-collected a total of 10 blood and 7 nasal swab samples over a 28-day observational period. For several infected participants, up to 4 additional weekly blood samples were collected to assess potential post-acute infection responses. 68 participants across 26 U.S. states completed the study (**Figure 1C**). A total of 691 out of 708 scheduled (98%) *home*RNA blood samples, 466 out of 476 scheduled (98%) nasal swabs, and 688 out of 708 administered (97%) online surveys were completed and returned (**Figure 2**). 22 participants were withdrawn; of which 13 were lost to follow-up and did not initiate study procedures (**Figure 2**). The median age was 38.0 years (IQR: 28.0 to 47.3; range: 20 to 74) (**Table 1**). The study enrolled participants from diverse demographics including individuals from understudied, underrepresented, and underreported (U3) populations; 71% female, 24% Hispanic, and a combined 25% of U3 minority (7% Asian; 6% Black, 12% mixed races) participants completed the study (**Table 1**).

**Table 1.** Demographics and clinical characteristics of participants who completed the study.

|  | All participants<br>(N=68) | high resolution immune profiling |  |
| --- | --- | --- | --- |
|  |  | COVID-19+ (N=8) | Uninfected (N=4) |
| Demographics |  |  |  |
| Sex, n/N (%) |  |  |  |
| Female | 48/68 (71) | 4/8 (50) | 1/4 (25) |
| Male | 20/68 (29) | 4/8 (50) | 3/4 (75) |
| Age, (years), median (Q1, Q3) | 38.0 (28.0, 47.3) | 38.0 (29.5, 44.0) | 40.0 (38.0, 43.0) |
| Ethnicity, n/N (%) |  |  |  |
| Hispanic | 16/68 (24) | 1/8 (13) | 1/4 (25) |
| Non-Hispanic | 52/68 (76) | 7/8 (88) | 3/4 (75) |
| Race, n/N (%) |  |  |  |
| Asian | 5/68 (7) | 0/8 (0) | 0/4 (0) |
| Black/African American | 4/68 (6) | 1/8 (13) | 0/4 (0) |
| White | 48/68 (71) | 7/8 (88) | 3/4 (75) |
| Mixed Race | 8/68 (12) | 0/8 (0) | 1/4 (25) |
| Unknown/Other | 3/68 (4) | 0/8 (0) | 0/4 (0) |
| Clinical characteristics |  |  |  |
| Weight (lbs.) |  |  |  |
| median (Q1, Q3) | 160 (141.3, 188.8) | 160 (147.8, 178.3) | 210 (183.8, 228.8) |
| Height (inches) |  |  |  |
| median (Q1, Q3) | 66.0 (64.0, 69.0) | 68.5 (66.3, 70.8) | 70.5 (68.5, 71.0) |
| COVID-19 vaccination, n/N (%) |  |  |  |
| Unvaccinated | 46/68 (68) | 2/8 (25) | 2/4 (50) |
| Vaccinated | 22/68 (32) | 6/8 (75) | 2/4 (50) |
| COVID-19+ infection timeline |  |  |  |
| Days, median (Q1, Q3; min, max) |  |  |  |
| post viral shedding | 1 (0, 2; -4, 6) | 0 (-1.5, 1; -4, 2) | na |
| post symptom onset | 0 (-1, 1, -5, 3) | -1 (-3, 0; -5, 3) | na |

**Figure 2.**
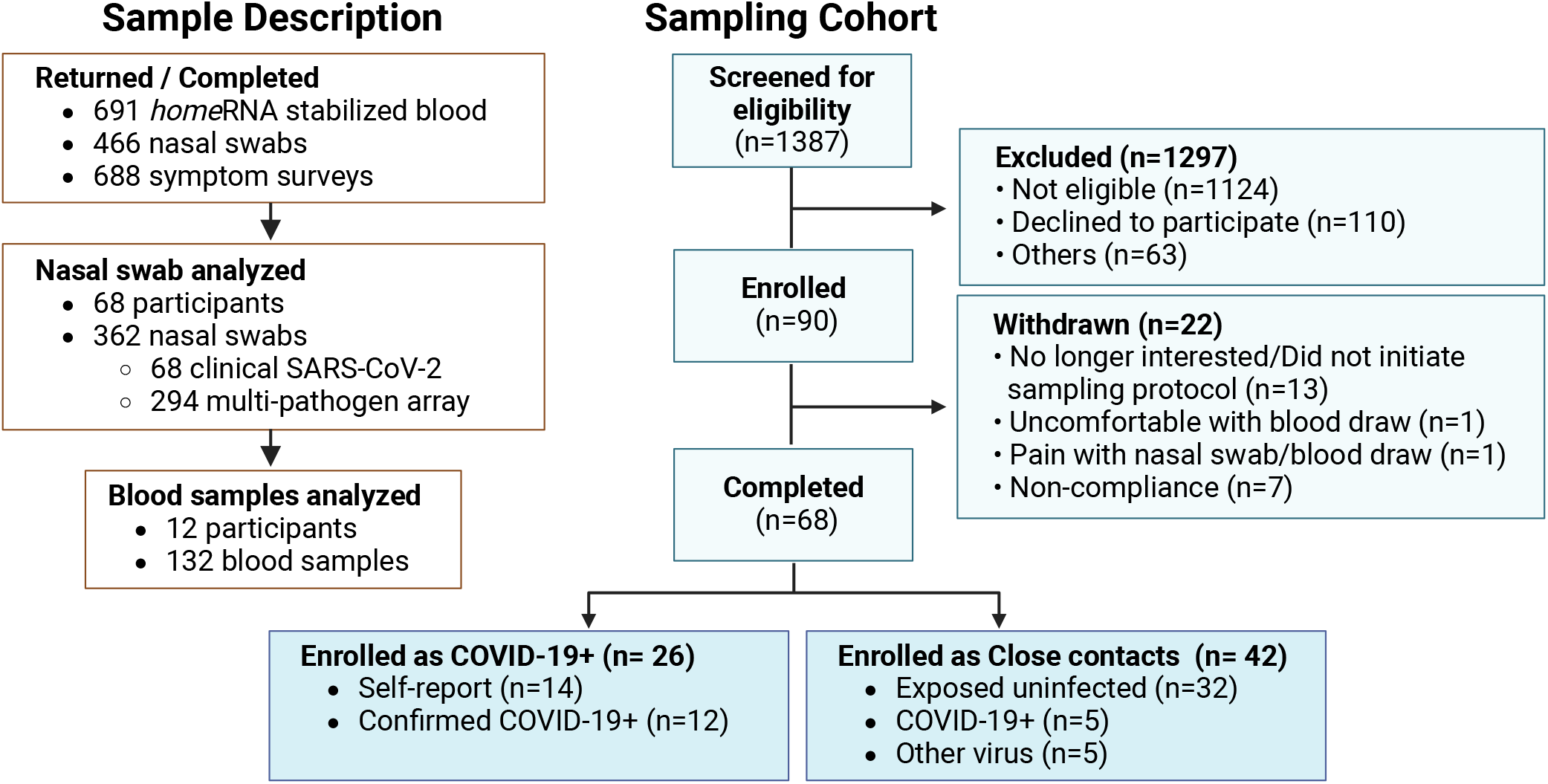
Participant and sample flow chart. Participants enrolled as COVID-19+ self-reported a recent SARS-CoV-2 positive test. Participants enrolled as close contacts had unknown infection status at the time of enrollment.

A total of 362 nasal swabs from all 68 participants were analyzed for respiratory pathogens (**Figure 2; Table S2**). 32% (22/68) of participants were positive for respiratory viruses in at least one swab (**Figure 2**). SARS-CoV-2 was associated with 77% (17/22) of infections. Among participants enrolled as exposed close contacts, 12% (5/42) were subsequently positive for SARS-CoV-2 (**Figure 3A**; **Figure S1A**). 16% (11/68) of participants reported recent SARS-CoV-2 infection but showed no virological evidence of infection in study swabs (COVID-19 probable) (**Figure S1A**). 32 exposed close contacts remained test-negative for all pathogens tested (exposed-uninfected) (**Figure 3A; Figure S1A**). Using a fully remote self-sampling study design, we were able to capture extremely early infection timepoints from community transmissions; on first sampling, the median days post viral shedding (days PVS; i.e., days after the first positive nasal swab) was 1 (range: -4 to 6 days) and the median days post symptom onset (days PSO; i.e., days after the first day of symptom onset as defined in the methods) was 0 (range: -5 to 3 days) (**Table 1**). Out of 17 confirmed SARS-CoV-2 infections, five participants (35%) were still presymptomatic when performed their first collection, of which, two were still in their pre-shedding phase (**Figure 3B**), providing us with a unique and unprecedented opportunity to characterize the earliest host immune kinetics to two major SARS-CoV-2 variants of concern (delta and omicron). Two SARS-CoV-2 infected participants remained asymptomatic throughout the observational window (**Figure 3B**).

**Figure 3.**
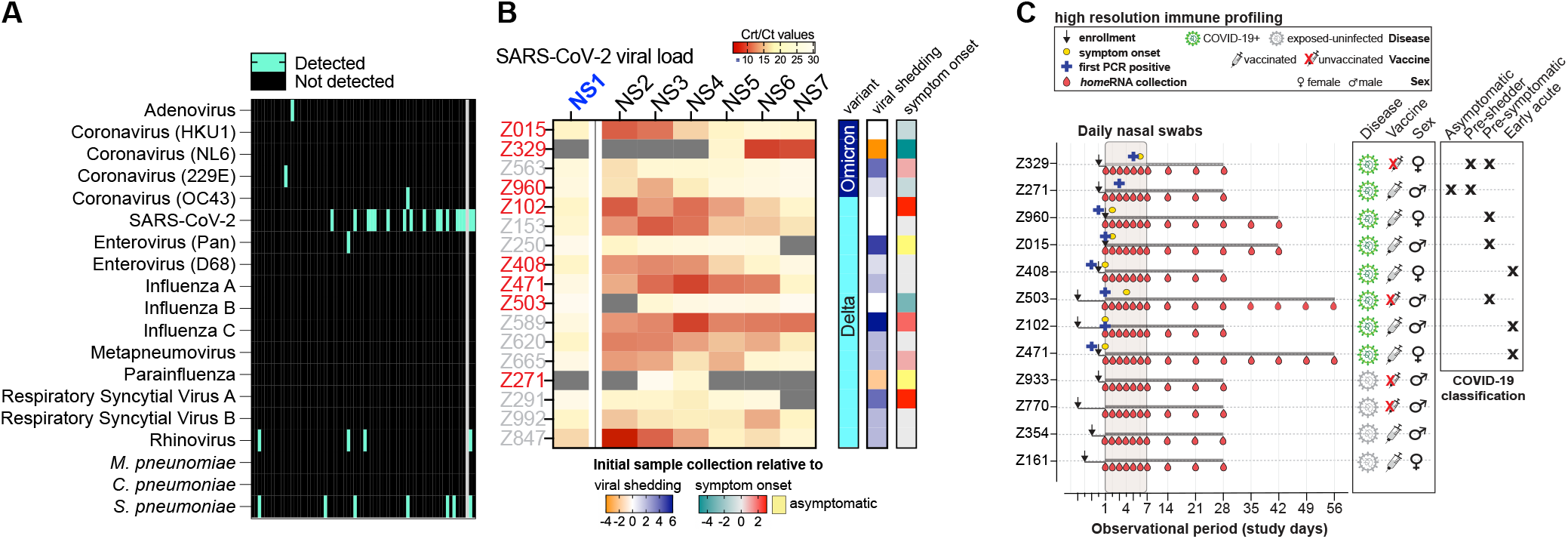
Virology status and classification of participants. A) Heatmap depicting pathogens detected in all 68 participants. Rows and columns represent pathogens and participants, respectively. 3-4 nasal swab per participant were screened for respiratory pathogens. Gray column denotes participant with a single nasal swab assessed for SARS-CoV-2 (not detected); this participant was not included in high-resolution immune profiling. **B)** Heatmap depicting SARS-CoV-2 viral load (n=17). Grey cells denote samples with undetectable viral load. Rows and columns represent participants and nasal swab samples, respectively. NS1 was subjected to clinical SARS-CoV-2 RT-PCR test (Ct values) and NS2-7 were subjected to a lab-developed multi-pathogen OpenArray assay (Crt values). Day 0 denotes onset of symptom (days PSO) and viral shedding (days PVS). Right annotation: viral shedding and symptom onset show a participant’s first sampling day relative to onset of viral shedding and symptom onset, respectively. Longitudinal blood samples from participants highlighted in red were subjected to high-resolution immune profiling. **C)** Clinical and sample characteristics of participants (8 COVID-19+ and 4 exposed-uninfected) selected for high-resolution immune profiling. Early immune kinetics were profiled from a total of 132 blood samples.

To evaluate the temporal dynamicity of host immune kinetics spanning the incubation, acute, and convalescent phases of SARS-CoV-2 infection, 92 longitudinal blood samples (10-14 timepoints/participant; -4 to 57 days PVS) from eight infected participants with the earliest infection samples were subjected to high-resolution immune profiling. (**Figure 1D**; **Figure 3C**). 785 genes associated with host susceptibility, interferon response, innate immune cell activation, adaptive immunity, and homeostasis (nCounter Host Response panel) were used to profile specific arms of the innate and adaptive immunity. GAMM were used to examine changes in gene expression for each disease status over time, using smooth functions of time (days PVS) to identify temporally dynamic genes within each disease group. Three disease groups were modeled: unexposed-uninfected (healthy), exposed-uninfected (exposed), and exposed-infected (COVID-19+). Using a 10% FDR, no dynamic genes were identified in either healthy or exposed participants, while 51% (324 significant and 73 trending) of the target genes were dynamic during SARS-CoV-2 infection (**Figure 4A**). Next, we performed hierarchical clustering of the Spearman’s rank correlation between every pair of gene-specific longitudinal trends (GAMM smooth fits) for the 324 significantly dynamic genes to identify gene clusters exhibiting similar longitudinal trends. Eleven gene clusters were identified and further reduced to four global temporal trends (**Figure 4B - 4E; Figure S2; Figure S3; supplemental data 4**). Temporal trends 1 and 2 consist of 69 early pre-shedding host response genes (**Figures 4B** and **4C; Figure S3**) with first peak expression estimated to occur prior to our observational window, followed in time by a vast number of early post-shedding genes (n=158) in temporal trend 3, most of which showed first peak expression within five days of viral shedding, and progressing to genes that peaked during mid-to-late acute phase of infection (n=97) in temporal trend 4 (**Figure 4D, Figure S3A**). It is important to note that, as with all analysis in this section, we are reporting our biological data as interesting observations afforded by this unique window into very early respiratory infection captured with high temporal frequency but from a limited sample size (n=8 participants with COVID-19); as described in the Discussion section, these observations need to be further validated with larger sample sizes before robust conclusions can be made.

**Figure 4.**
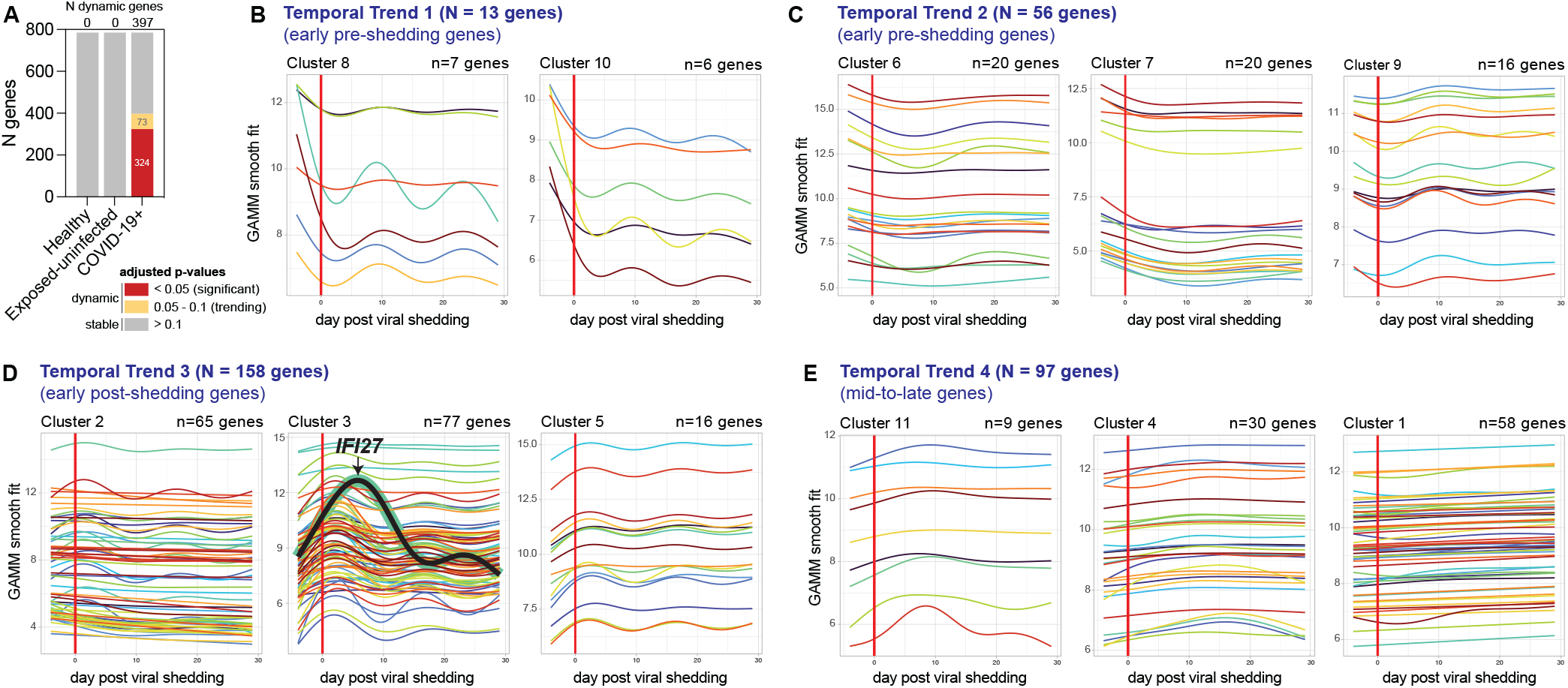
Dynamic genes identified during SAR-CoV-2 infection. A) Number of dynamic genes identified through smooth function of time (day PVS) in healthy, exposed-uninfected, and COVID-19+ participants. (B-E) GAMM smooth fits of dynamic genes derived from COVID-19+ participants (n=8) and clustered by their temporal profiles (see **Figure S2** for heatmap showing clustering of dynamic genes by temporal trends). Clusters depicting similar longitudinal trends were further reduced to four global temporal trends (B-E). Red vertical line denotes onset of viral shedding (see **Figure S3C** and **Table S3** for gene membership within each cluster).

Figure 5. illustrates the dynamics of gene expression in distinct pre- and post-shedding gene clusters across different participants. Marked induction of pre-shedding genes (Clusters 6, 7, 8, and 10) was observed in participants (Z329 and Z271) who initiated collection during this phase irrespective of symptom outcomes (**Figure 5**). During the pre-shedding phase, induction of genes associated with various innate and adaptive immunity such as broad immune cell mobilization (e.g. granulocytes, leukocytes, myeloid leukocytes, lymphocytes), regulation of general stress and inflammatory responses (e.g. MAP kinase cascade signaling, chemokine signaling, cytokine production, oxidative stress), cytotoxic processes (e.g. cell killing, apoptotic signaling), and T cell mediated immunity (e.g. αβ T cell activation, lymphocyte differentiation and migration) were observed (**Figure S4; Figure S5**). Peak expression of these genes occurred prior to the earliest timepoint (D-4), followed by a rapid loss of response by the first onset of viral shedding (D0). Thus, we were unable to evaluate whether these signatures are shared across other infected participants who initiated sampling after onset of viral shedding. However, a trend towards peak before viral shedding can be observed (**Figure 5**; green arrows) in two additional infected participants (Z102 and Z503) with early D0 timepoint samples.

**Figure 5.**
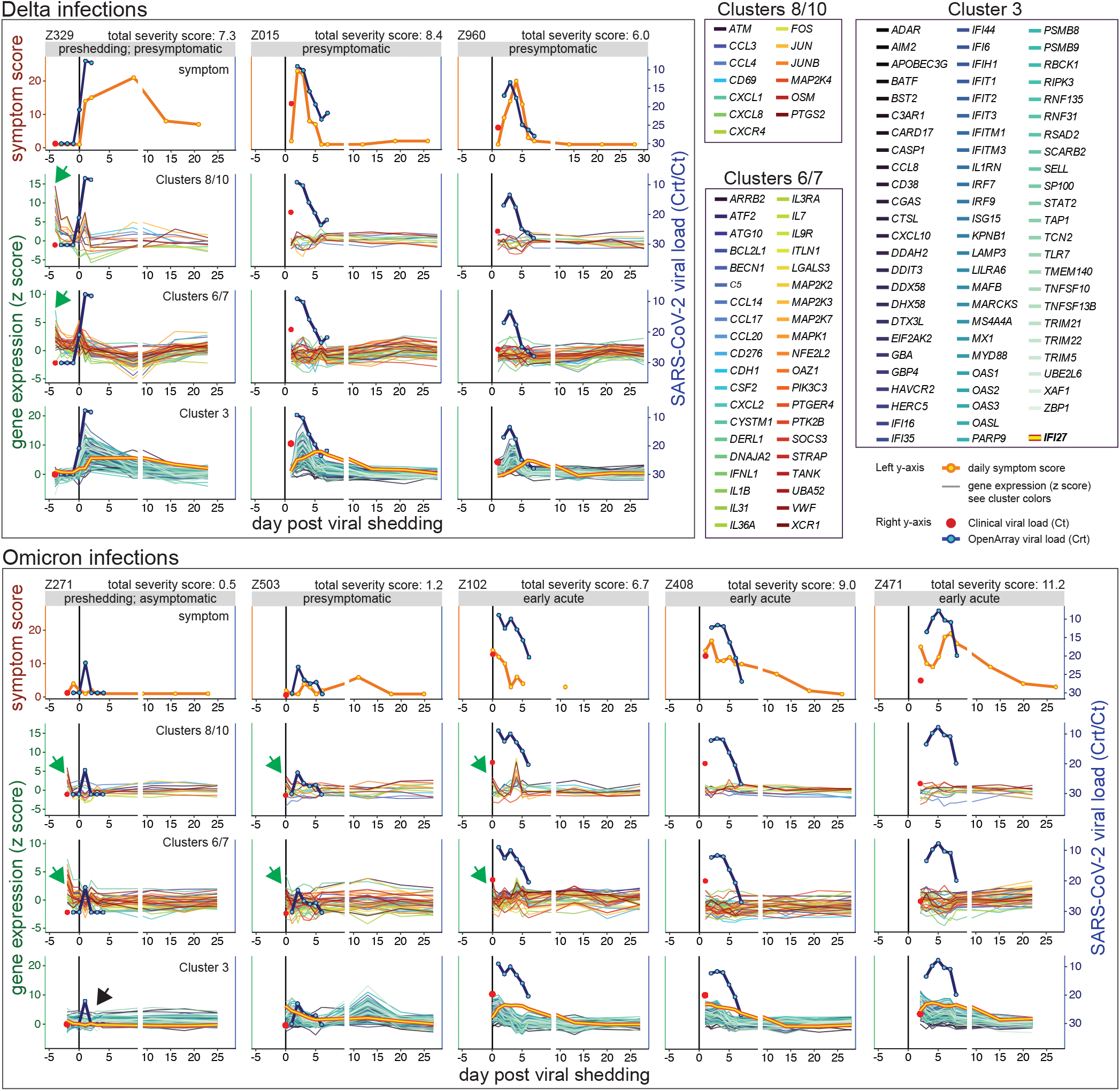
Detailed immune kinetics of early pre- and post-shedding genes in individual participants. Participants were grouped by SARS-CoV-2 variants. Within each variant group, columns represent individual participants; rows represent daily symptom burden of individual participants (row 1) and expression of early pre-shedding (clusters 8/10 and clusters 6/7, rows 2 and 3, respectively) and post-shedding (cluster 3, row 4) genes. Within each plot, black vertical lines denote onset of viral shedding. Symptom burden and gene expression are plotted on the left y-axis. Viral load is plotted on the right y-axis. x-axis denotes day post start of viral shedding. Viral load measured using clinical SARS-CoV-2 RT-PCR tests (first sample for each participant) are reported as Ct values and plotted as a single red point while viral load measured using the lab-developed multi-pathogen OpenArray assay are reported as Crt values and plotted as lines. Gene expression z scores were computed using the mean and standard deviations measured from unexposed-healthy controls. Green arrows denote increased pre-shedding signatures in two participants with sampling timepoints at 0 day PVS (Z503 and Z102) in addition to the two participants who initiated sampling in the preshedding phase (Z329 and Z271). Black arrow denotes lack of induction response in one asymptomatically infected participant. *IFI27* immune kinetics (time-trend 3) highlighted in orange with red contours.

Active viral replication and onset of viral shedding led to induction and further augmentation of early post-shedding genes (Clusters 2, 3, and 5) associated with a broad repertoire of innate and adaptive signaling pathways, including antiviral defenses that modulate viral entry into host cells or containment of infected cells (phagocytosis, programmed cell death, cell killing), interleukin production, Type I/II interferon responses, and T cell activation, differentiation, and proliferation signatures (**Figure S4**; **Figure S6**). During this period, increased symptom burden along with robust expression of interferon-stimulated genes (ISGs; Cluster 3) was observed across all infected, symptomatic participants (**Figure 5**; **Figure S4**). The ISG response depicted tight co-regulation marked by a homogenous temporal trend (**Figure 5**). However, detailed immune kinetics during early infection afforded through frequent daily measurements uncovered the decoupling of *IFI27* kinetics from other ISGs, marked by delayed and prolonged expression in several symptomatic participants (**Figure 5**). Although *IFI27* was recently identified as a biomarker for early presymptomatic screening of SARS-CoV-2 infection, validation studies incorporating prominent presymptomatic genes identified in this work may improve the detection of presymptomatic cases^13^. A two-day low viral shedding period coupled with lack of ISG response (black arrow) was observed in one asymptomatic infection (Z271; **Figure 5**). Furthermore, assessment of both SARS-CoV-2 viral kinetics and self-reported symptom burden depicted a striking alignment between ISG response in the periphery and viral kinetics at the site of infection (**Figure 5**), highlighting the complex associations and interactions between systemic (peripheral) host immunity and localized (mucosal) viral factors.

GAMM smooth function fits obtained through daily measurements enabled granular observations on the timing when an immune response first occurred, allowing us to characterize the initial response of distinct immune mechanisms within two host antiviral defense pathways involved in interferon signaling (Type I and II) and antigen presentation (MHC Class I and II) (**Figure 6**). For both Type I and Type II IFN pathways, we observed induction of transcriptional regulators such as subunits of the AP-1 transcriptional complex (*FOS*/*JUN*) and MAP kinase signaling cascade (*MAPK1*) occurred prior to viral shedding (**Figure 6A**). Upon onset of viral shedding, an earlier IFN*γ* response followed by IFNα/β signaling can be observed in the periphery as evidenced by earlier peak expression of IFN*γ* response genes. (**Figure 6A**). Both *STAT2* and *IRF9* required for transcriptional activation of Type I-responsive ISGs first peaked at 1.8- and 2.1-day PVS respectively, while *JAK2* and *IRF1*, involved in transcriptional activation of IFNγ-responsive genes peaked at 1.3- and -0.4-day PVS respectively (**Figure S7; Figure 6C; Figure 6D**). Peak expression for many IFNα/β genes occurred within a narrow temporal window during infection, further corroborating the tight co-regulation of this pathway during infection (**Figure 6A**). Both MHC class I and II antigen presentation are crucial to elicit robust T cell responses to infection. For example, MHC class I-mediated presentation of viral peptides to CD8+ T cells is crucial for killing of virus-infected cells and effective pathogen control. Similar to the early IFN response, we found that expression signatures associated with MHC class I antigen presentation were induced in the periphery during early infection with most genes peaking within 5 days PVS, followed in time by MHC Class II signatures (**Figure 6B**), including robust expression of genes directly involved in antigen processing (*TAP1, CTSL*), ubiquitination (*UBE2L6, HERC5*), proteosome degradation (*PSMB9*), and peptide loading (*HLA-A, HLA-B, HLA-E*) (**Figure 6E**).

**Figure 6.**
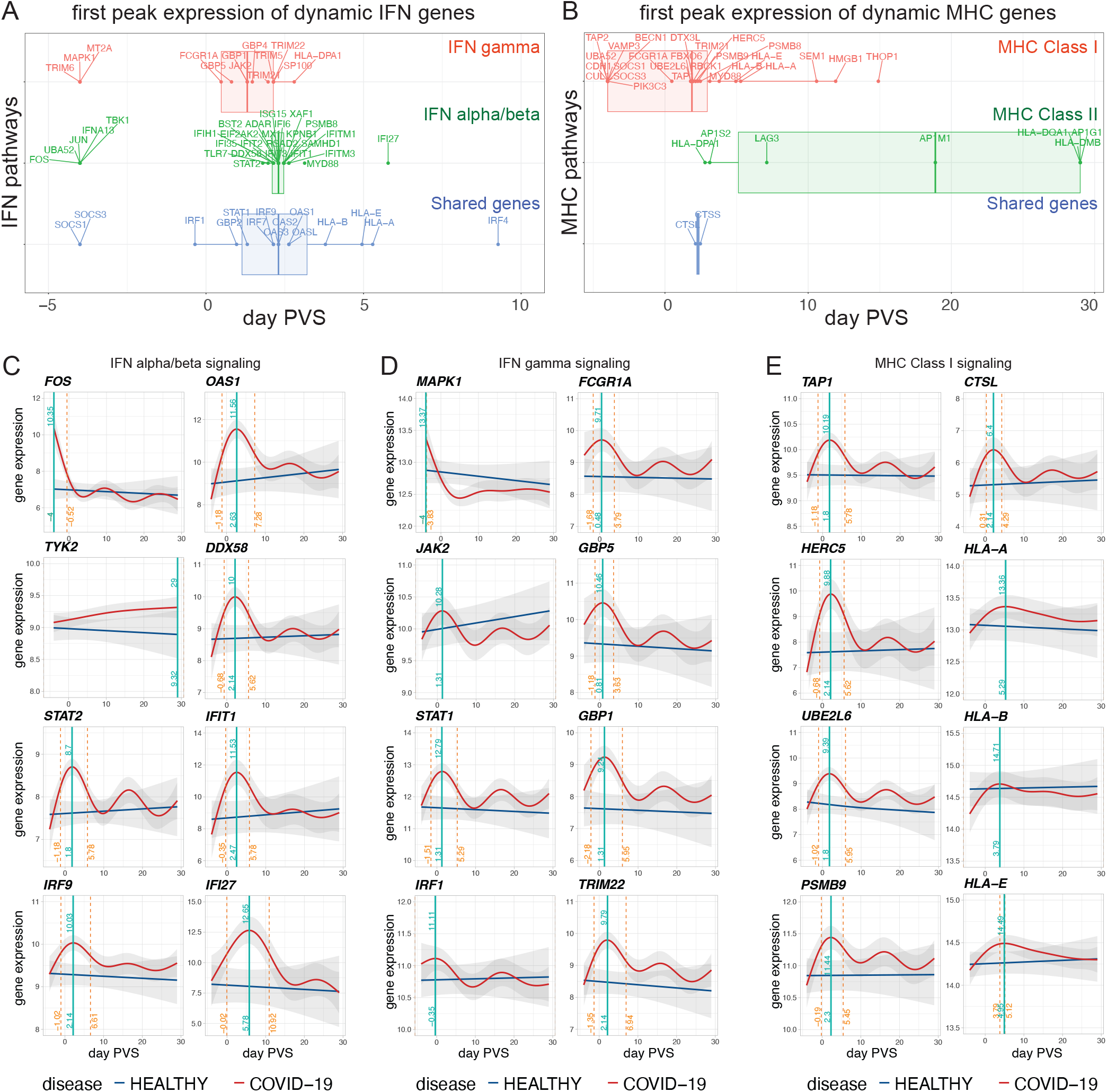
Immune kinetics from self-collected longitudinal samples revealed early activation of IFNγ signaling and MHC class I dependent antigen presentation during COVID-19 infection. Boxplots depicting the day PVS corresponding to the first peak expression derived from GAMM smooth fits of genes within the A) interferon signaling and B) MHC antigen presentation pathways. (C-E) GAMM smooth fit depicting immune kinetics of select D) IFNα/β, E) IFNγ, and F) MHC class I antigen presentation genes in healthy and COVID-19+ participants. For IFN pathway genes (C and D), upstream regulators required for activation of ISRE (IFNα/β) and GAS (IFNγ)-dependent ISGs shown in **Figure S7** and several key ISGs within that pathway were depicted. For MHC Class I genes (E), direct molecular components of the MHC Class I antigen processing and presentation pathway were depicted. For each gene, the duration (day PVS) between the vertical orange dotted lines denote the time period during infection for when the smooth fit for COVID-19+ is estimated to differ from that of healthy individuals, as defined by the lack of overlap between the 95% confidence interval predicted for each disease group. For each gene, the peak expression of the first response is marked by a vertical green line. A value of -4 was assigned to genes whose peak expression was estimated to occur prior to the observational window of (D-4).

Next, we assessed if baseline blood transcriptional signatures (at the time of exposure) are associated with protection from infection in exposed-uninfected individuals. Using linear mixed models (LMM), we first found 261 genes showing significant differential response in the three disease groups (healthy, exposed-uninfected, and infected participants); we then contrasted pairs of disease groups for differential response: “exposed-uninfected vs healthy”, “COVID-19+ vs healthy”, and “COVID-19+ vs exposed-uninfected” (**Figure S8**). Although no genes were found to be temporally dynamic in exposed-uninfected and healthy participants (GAMM analysis; **Figure 4A**), a total of 108 genes were differentially expressed between exposed-uninfected and healthy participants at baseline (**Figure S8A**). Genes overexpressed in exposed-uninfected participants relative to their exposed-infected counterparts can potentially play a role in host protection against acquisition of infection. GSEA performed using ranked LMM estimates (see statistical methods) from “COVID-19+ vs exposed-uninfected” suggested that neutrophil degranulation, bacterial infection, and antimicrobial peptides pathway genes were overexpressed in exposed-uninfected participants (**Figure 7, Figure S8B, Figure S9A**) while known antiviral mechanisms including IFN*γ* signaling were overexpressed in COVID-19+ participants (**Figure 7B Top Panel, Figure S8B, Figure S9B**).

**Figure 7.**
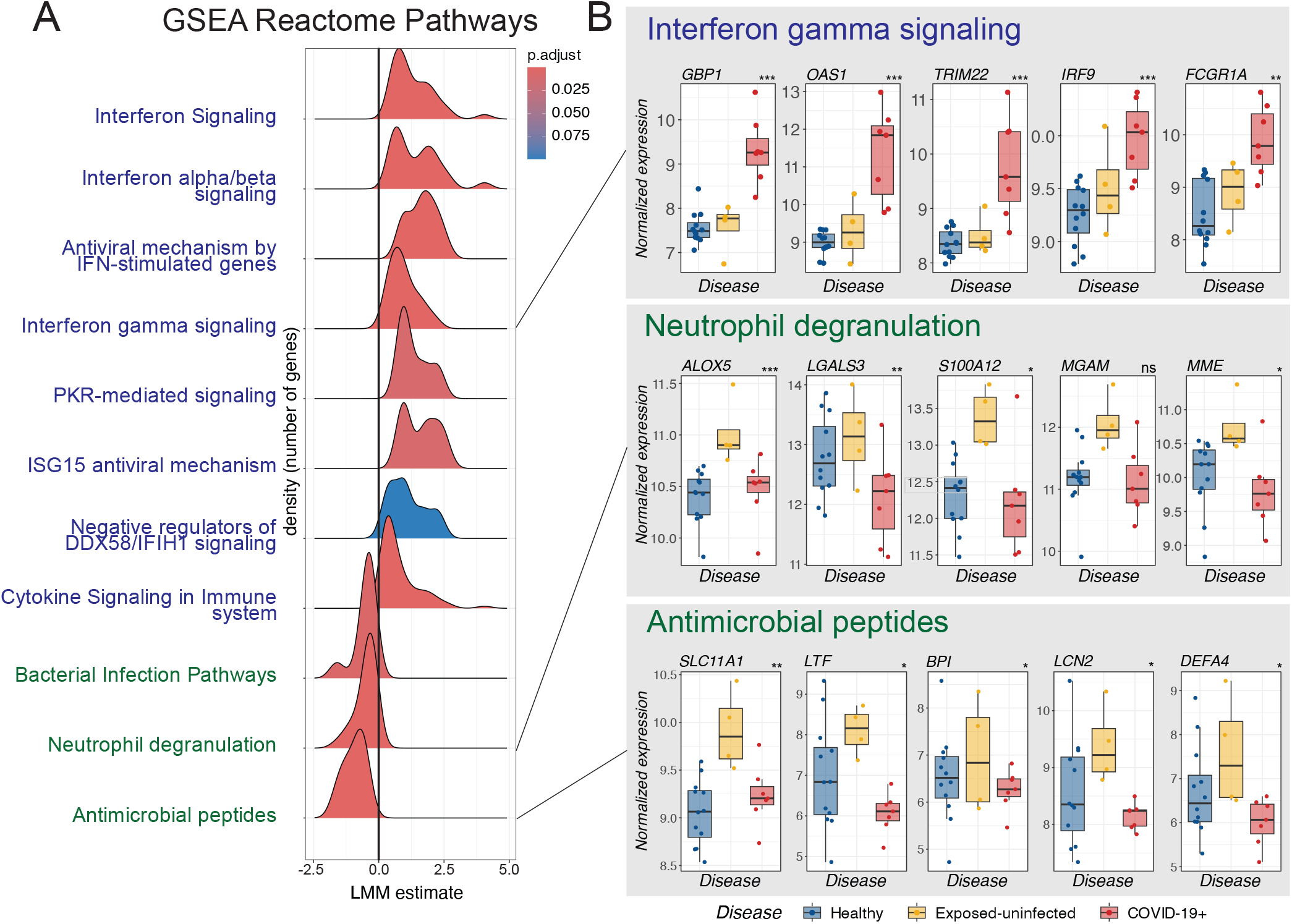
Increased expression of neutrophil degranulation, host defenses to bacterial infection, and antimicrobial peptide-encoding genes in exposed-uninfected individuals. A) Ridgeline plot summarizes the results from GSEA analysis. Each row shows the distribution (density plot) of resulting LMM estimates of the average difference between gene expression measured in COVID-19+ and in exposed-uninfected (reference) participants for genes within each significant GSEA pathway. GSEA was performed using a ranked list of LMM estimates obtained from “COVID-19+ vs exposed uninfected” pairwise comparisons. Negative LMM estimates denote increased expression in exposed-uninfected participants while positive LMM estimates denote increase expression in COVID-19+ participants. B) Boxplots showing Day 1 expression of IFNγ, neutrophil degranulation, and antimicrobial peptide pathways in healthy, exposed, and COVID-19+ participants. Statistical significance between healthy, exposed, and COVID-19+ disease groups were determined using LMM (see methods for details). P-values were adjusted for multiple testing correction using the BH procedure and significance between disease groups were labeled (top right) as follows: p >0.05 (ns), p ≤0.05 (*), p ≤0.01 (**), p ≤0.001 (***).

## DISCUSSION

Early dynamics of host immunity following pathogen exposure can offer insights into pathways that confer susceptibility or protection against infection, or against symptoms despite infection. The earliest host transcriptional responses to viral infections can be rapid and transient ^21-23^, making them hard to capture in natural infections when exposure timelines cannot be controlled. In-person venipuncture still presents a logistical barrier to conducting longitudinal studies requiring frequent blood collections; high-frequency immune profiling of early blood transcriptional response is largely limited to controlled human challenge studies and there is currently an unmet need for a study framework that enables conduct of such studies in the natural field setting and in natural exposures to infections acquired through community transmission.

Here, we present a framework for conducting fully remote longitudinal studies with decentralized sampling (self-collection of blood for transcriptomics and nasal swabs) to characterize the dynamicity of a rapidly evolving immune state during early stages of ARIs and report its first real-world application and feasibility in profiling extremely early host immunity to natural respiratory infections. To the best of our knowledge, this is the first prospective longitudinal nationwide study to perform high-frequency intra-individual measurements spanning the pre-shedding-to-convalescence phases of infection. Using home self-collection tools, we synced daily kinetics of viral load and host blood transcriptional response in natural infections.

A key strength of the study lies in its decentralized self-sampling to enable rapid dissemination of collection kits to participants over wide geographical areas, large-scale recruitment across diverse communities throughout the United States, and frequent biomarker measurements from the blood responses at early or dynamic timepoints (**Figure 1C**). For highly contagious infections, especially during public health emergencies such as the COVID-19 pandemic, decentralized self-sampling enables us to observe the natural history of disease while maintaining isolation of infected participants and eliminating exposure risks associated with in-person studies during early disease when participants are most infectious.

Given that duration of the incubation period varies significantly between individuals and can be as short as several days, minimizing the interval between screening of eligible individuals and first sample collection was integral to capturing exposed close contacts who were still in their presymptomatic phase. 35% of infected participants were still presymptomatic on their first day of sampling, demonstrating feasibility of this study framework to quickly identify and capture early infection timepoints. In fact, a small but interesting subset of infected participants initiated sampling prior to detectable nasal viral shedding and provided a unique opportunity to obtain high-frequency measurements of the earliest transcriptional changes in the periphery. These pre-shedding observations are extremely hard-to-capture in natural infections and mostly observed within the setting of human challenge studies. Future studies with targeted recruitment in high-risk settings coupled with same-day delivery of study kits to exposed close contacts could further enrich for presymptomatic and pre-shedding timepoint samples.

Detailed temporal trajectories of 785 innate and acquired immune genes along with temporally aligned nasal viral load profiled for each individual participant led us to several key findings. While both exposed-uninfected and healthy participants showed complete absence of dynamic genes, a large repertoire of dynamic immune genes were identified during COVID-19 infection, enabling us to profile detailed immune kinetics during early infection (**Figure 4A**). Notably, robust expression of many ISGs known to drive the early inflammatory response to respiratory viruses were lacking during the pre-shedding phase of infection (**Figure 5, Figure S3, Figure S4, Figure S6**). Instead, we identified a group of transient early response genes that were induced in both symptomatic and asymptomatic infections during pre-shedding timepoints (**Figure 4, Figure 5**) and found that the early transcriptional response in the periphery consists of genes associated with broad activation and mobilization of leukocyte and T cells, chemokine signaling, various stress response pathways, cytotoxicity, and regulation of apoptotic signaling (**Figure S5**). The rapid loss of this peripheral response at onset of nasal viral shedding coupled with marked chemotaxis signatures suggest rapid immune cell recruitment to the site of infection^24^. Onset of viral shedding is marked by robust expression of ISGs, with the exception of one asymptomatic participant, who showed complete absence of IFN response (**Figure 5, Figure S4**). Despite lack of ISG response, expression of pre-shedding signatures can still be observed in asymptomatic infection (**Figure 5**). The absence of IFN response in asymptomatic Omicron infection corroborates recent findings from a SARS-CoV-2 challenge study using a pre-Alpha SARS-CoV-2 challenge strain^24^. Given the small sample size of our study, it will be important to determine if our findings continue to be corroborated by additional future studies by us and others.

Second, we demonstrated that highly resolved immune kinetics can be used to characterize differences in the timing of activation between distinct immune arms during dynamic stages of infection; using two major antiviral pathways, we showed that IFN*γ* signaling precedes IFNα/β signaling (**Figure 6A**). Furthermore, early MHC class I dependent antigen presentation signatures were present in the periphery (**Figure 6B**), including expression of direct components of the antigen processing and cross-presentation mechanism further supporting the presence of early mucosal response signatures in the blood (**Figure 6E**). Taken together, these observations suggests that T cell immunity and T cell-mediated cell killing is present very early in infection and can be detected in the blood (**Figure 6B, Figure S5**). With the exception of *IFI27*, IFNα/β signaling genes portrayed highly homogenous kinetics throughout the observational window and strong transcriptional co-regulation (**Figure 4B, Figure 5**). Decoupling of *IFI27* expression kinetics from kinetics of other ISGs marked by a prolonged induction period and delayed peak expression at 5.8 days PVS suggests a distinct transcriptional network or molecular machinery governing *IFI27*-associated antiviral mechanism, laying the foundation to further test this hypothesis in future studies with larger sample sizes.

Third, comparison between exposed-infected vs exposed-uninfected participants led us to identify signatures that may play a role in host susceptibility to infection. Abortive subclinical infections have been previously observed in individuals exposed to SARS-CoV-2^24,25^. In human viral challenge studies, only a subset of experimentally challenged participants developed clinical infections, suggesting the role for preexisting immunity (cross-reactive or pathogen-specific) in exposed individuals who did not become infected or develop clinical disease^8,15,24^. In our study, elevated expression of antimicrobial peptides was observed in exposed-uninfected individuals relative to infected and unexposed-healthy participants (**Figure 7**). The antimicrobial properties that confer protective immunity against various bacterial, fungal, and viral pathogens are multifaceted ranging from indirect immunomodulation of the inflammatory cascades to direct nutritional immunity through metal sequestration and binding of microbial siderophores, an iron-acquisition small molecule produced by many bacteria and fungi. During infections with SARS-CoV-2, defensins have been demonstrated to restrict viral entry in *in vitro* studies^26^. Due to their longstanding reputation as key innate defenders, antimicrobial peptides were quickly proposed in therapeutic applications against SARS-CoV-2^27^ based upon antiviral efficacy observed from *in vitro* cellular assays. Our findings provide evidence from human studies of the potential protective role of these peptides against SARS-CoV-2. They could be validated in a larger cohort, including for SARS-CoV-2 and other ARIs, using the decentralized study design introduced in this work.

There are several limitations to the study. First, our cohort was small in size and included a limited number of presymptomatic and pre-shedding timepoints. Despite this limitation, the relatively frequent intra-individual measurements (daily measurements in the first week) coupled with longitudinal GAMM analyses enabled detection of early pre- and post-shedding signatures (**Figure 4**). Second, unsupervised home self-sampling methodologies are inherently susceptible to user-associated variations (e.g. user skills, transit duration, and post-collection storage conditions) that can impact sensitive gene expression readouts. However, the reliability of Tasso-based self-collection devices for antibody measurements and pathogen testing (e.g. HIV, Syphilis, CMV) have been demonstrated by other groups ^28-30^. In this and prior studies from our group, the lack of dynamic genes in both healthy and exposed-uninfected individuals demonstrated that the study and analysis framework remained robust to user-associated variations^19^. Lastly, the use of a targeted gene panel as opposed to genome-wide transcriptomics in addition to lack of evaluation of the localized host response in the airway limits a comprehensive measurement of the entire transcriptome and of the immune response at the site of infection during early infection. Future studies using genome-wide transcriptome coupled with evaluation of host mucosal response will enable to correlate blood and mucosal responses and provide a more comprehensive understanding of the host immunity upon pathogen exposure.

In sum, we successfully demonstrated the feasibility of decentralized sampling using home self-collection tools to capture early, transient changes in host response soon after viral exposure in natural infections at high temporal resolution comparable to human challenge studies. Overarchingly, our study provides a technological and study framework—using *home*RNA—to investigate detailed kinetics of host immunity to not just ARIs, but various infectious and non-infectious diseases with dynamic and rapidly evolving immune states, such as those characterized by recurrences or relapses (e.g., autoimmune disorders and malignancies). Most importantly, providing the autonomy of completing study procedures from a home setting to study participants themselves enables conduct of large-scale population-wide mechanistic studies across diverse communities. By reducing study burden, decentralized sampling has significant potential to augment enrollment of participants most affected by logistical barriers, particularly individuals from underreported, underserved, and understudied communities.

## Data Availability

All data produced in the present study and R scripts developed to perform analyses and visualizations are available upon reasonable request from the authors.

## Author contributions

Conceptualization: FYL, KA, ABT, EB, AW

Methodology: FYL, HGL, AD, LS

Investigation: FYL, HGL, AD, LS

Analysis and visualization: FYL, HGL, SYK, TVN, OH

Funding acquisition: ABT, AW

Project administration: FYL, GH, AD, MT, KA, HGL, ABT

Supervision: ABT, EB, AW, FYL

Writing – original draft: FYL, HGL, SYK, ABT, EB, AW

Writing – review & editing: FYL, HGL, SYK, TVN, LS, MB, JTS, OH, AW, ABT, EB

We would like to acknowledge Dr. Kimberly Dill-McFarland at the Department of Medicine, University of Washington for discussions surrounding bioinformatic analysis. We would like to thank Caitlin Wolf, Tessa Wright, and Devon McDonald from the Husky Coronavirus Testing Program for participant recruitment and outreach. We would also like to thank the study participants. The REDCap instance used is supported by the Institute of Translational Health Sciences, which is funded by the National Center for Advancing Translational Sciences of the National Institutes of Health under award number UL1TR002319.

## Data and material availability

All data are available in the main text or the supplementary materials. nCounter gene expression data available as normalized counts in supplemental data 1. R scripts developed to perform analyses and visualizations are available upon request from the co-senior authors.

