## Supplemental File for "*home*RNA self-blood collection enables high-frequency temporal profiling of presymptomatic host immune kinetics to respiratory viral infection: a prospective cohort study"

*Co-senior authors:

### Supplemental Methods

#### *home*RNA blood collection

**Sampling kit assembly and device fabrication:** Tasso-SST blood collection devices were purchased from Tasso, Inc. The stabilizer tube components were injection molded out of polycarbonate (PC: Makrolon 2407) by Protolabs, Inc (Maple Plain, MN). Fabrication of the stabilizer tube and components of the *home*RNA blood collection kit has been previously described[1].

**Blood collection**: Blood collection was performed at the residential homes or other temporary locations (if traveling during study) on pre-scheduled sampling days. Detailed description of the *home*RNA collection procedures has been previously published[2]. Briefly, after sanitizing and warming the collection site (upper arm), Tasso-SST blood collection device was adhered, activated, and up to 0.5 mL of liquid capillary blood was collected. Immediately after collection, participants connected the blood collection tube to the stabilizer tube containing RNAlater and to mix and complete stabilization (**Figure 1B**). Stabilized blood was packaged according to the return instructions, stored at ambient temperature, and returned to the lab via overnight courier services. Samples collected on weekdays were mailed back to the University of Washington within 24 hours of collection using Next Day Courier Service (overnight shipping) and delivered directly to a secure -20ºC freezer. When collection was scheduled on a weekend, participants were asked to store stabilized samples in a cool location until the first available courier pick up service the following Monday. Samples were transferred to -80ºC for long term storage until further sample processing.

#### Respiratory specimen collection, viral load kinetics, and SARS-CoV-2 viral sequencing

Participants were asked to collect nasal swab samples from both nostrils using a sterile polyester-flocked swab (Puritan PurFlock Ultra; #25-3306-U) and transfer swabs into universal transport medium (Copan Diagnostics) immediately after collection. The first nasal swab (NS1) was returned to a CLIA-certified lab for SARS-CoV-2 RT-PCR testing and the remaining swabs (NS2 - NS7) were returned to the University of Washington for multi-pathogen array analysis. Nasal swabs were delivered directly to a secure -20ºC freezer and transferred to -80ºC for long term storage. Nucleic acid extraction and pathogen quantification using the Multi-pathogen OpenArray platform for non-clinical testing of respiratory pathogens was performed as previously described[2]. For each participant, 3-4 nasal swabs were tested for SARS-CoV-2 and 18 common respiratory pathogens as previously described.

#### Online surveys

All surveys were administered online via REDCap[3]. A Welcome Survey was administered immediately upon screening and consenting. The survey collected information on recent SARS-CoV-2 test, COVID-19 vaccination history, clinical traits (age, sex, weight), ethnicity and race, living arrangements, and pre-existing health conditions. Daily Use Surveys were administered on each scheduled collection timepoints and collected symptom burden information and usage parameters associated with the self-sampling devices. Two general surveys intended to query general user experience pertaining to the decentralized study framework such as timeliness of courier services and convenience of the specimen return logistics were administered at the end of week 1 and again, at the completion of all study procedures.

#### Gene expression analysis

**RNA isolation, cleanup, and concentration**

Total RNA was isolated using the Ribopure™ Blood RNA Isolation Kit (Thermo Fisher #AM1928) according to the manufacturer-recommended alternate protocol for enrichment of small RNAs (e.g. miRNA, tRNA, 5S/5.8SrRNAs). Briefly, stabilized blood cells were pelleted, lysed using 800 µL of lysis solution and 50 µL of acetic acid, and RNA was extracted using 500 µL of acid-phenol:chloroform (PCI). The RNA-containing aqueous phase was collected and denatured with 1 mL Denaturation Solution (Ambion #AM8540G) and 1.25 v/v of 100% ethanol. Nuclease-free water was added in 300 µL increments until solution turned clear. For the final purification of total RNA enriched with small RNAs, extracted RNA was bound to a silica filter cartridge, washed once with 70:30 (v/v) ethanol: denaturation, followed by two washes of the final wash solution (80% ethanol/50mM sodium chloride), and eluted twice in 100 µL of total elution volume. RNA yield was quantified on the Cytation5 Take3 plate. RNA quantification for nCounter gene expression analysis was performed on the Qubit 4 fluorometer using the Qubit RNA HS Assay Kit (Thermo Fisher). RNA quality was measured on the Bioanalyzer 2100 (Agilent Technologies). RIN values for RNA samples with < 5 ng/µL were assayed using the RNA 6000 Pico Kit (Agilent Technologies #5067-1513) and > 5 ng/µL were assayed using RNA 6000 Nano kit (Agilent Technologies #5067-1511). Low concentration samples were concentrated using Monarch RNA Cleanup Kit 10 µg (NEB #T2030) according to manufacturer's guidelines. RNA samples were stored at -80 ºC until ready for nCounter gene expression analysis.

**nCounter gene expression analysis:** The nCounter Pro Analysis System (nanoString) was used to perform direct detection and digital counting of native RNA transcripts. 50-120 ng of total RNA from each participant sample were hybridized to the nCounter Host Response Panel codeset version v1.1 (nanoString) to generate the expression dataset. The nCounter Host

Response codeset targets 773 genes associated with the immune response to infectious disease along with 12 candidate reference (housekeeping) genes. The target-probe hybrids were immobilized on a cartridge, aligned, and digitally counted on the nCounter Pro digital analyzer.

**nCounter data quality control and normalization:** Raw nCounter expression counts were normalized using the nSolver$^{TM}$ software (nanoString). Quality control (QC) and normalization procedures were performed as previously described[2]. All samples passed QC metrics for imaging (> 75% field of view), binding density (0.1 – 2.25 spots/square micron), and positive control linearity (> 0.95). Samples were subjected to i) positive control normalization and ii) codeset content normalization. Five reference genes (*GUSB, HRPT1, MRPS7, NMT1, PGK1*) were used in codeset content normalization. Normalized expression counts are provided in supplemental data 1. Normalized counts were transformed to trimmed mean of means (TMM) log2 counts per million for linear mixed effect modeling and generalized additive mixed modeling.

**Other data visualization:** Volcano plots and box plots were generated in R Statistical Software (v4.2.1, R Core Team 2022)[4]. Study workflow and conceptual diagrams were constructed in Biorender and figures were compiled in Adobe illustrator.

**References used for statistical analyses**

Gene expression was modeled as trimmed mean of means (TMM) normalized $\log_2$ counts per million[5]. Generalized additive mixed models (GAMM) were used to evaluate associations between gene expression and disease status (healthy, exposed-uninfected, and COVID-19 positive) over time[6]. GAMM analyses with thin plate regression splines with 6 basis dimensions were carried out using the gamm4 R package[7]. P-values were adjusted for multiple comparisons by controlling the false discovery rate (FDR) using the Benjamini-Hochberg (BH) procedure[8].

Linear mixed effect models (LMM) within the kimma R package were used to examine the influence of disease status on gene expression during the early stages of the disease[9]. Gene-level quality weights were calculated using voomWithQualityWeights in limma R package[10] and incorporated into the model to account for gene-level variability between different observations[11].

Gene ontology overrepresentation analyses (GO-ORA) were performed using the enrichGO() function of the clusterProfiler R package[12,13]. For parent term analyses, semantic similarity between pairs of GO terms were calculated and a matrix of dissimilarities was computed using the calculateSimMatrix() function of the rrvgo package[14]. Geneset enrichment analysis (GSEA) were performed using gsePathway() function of the ReactomePA R Package[15].

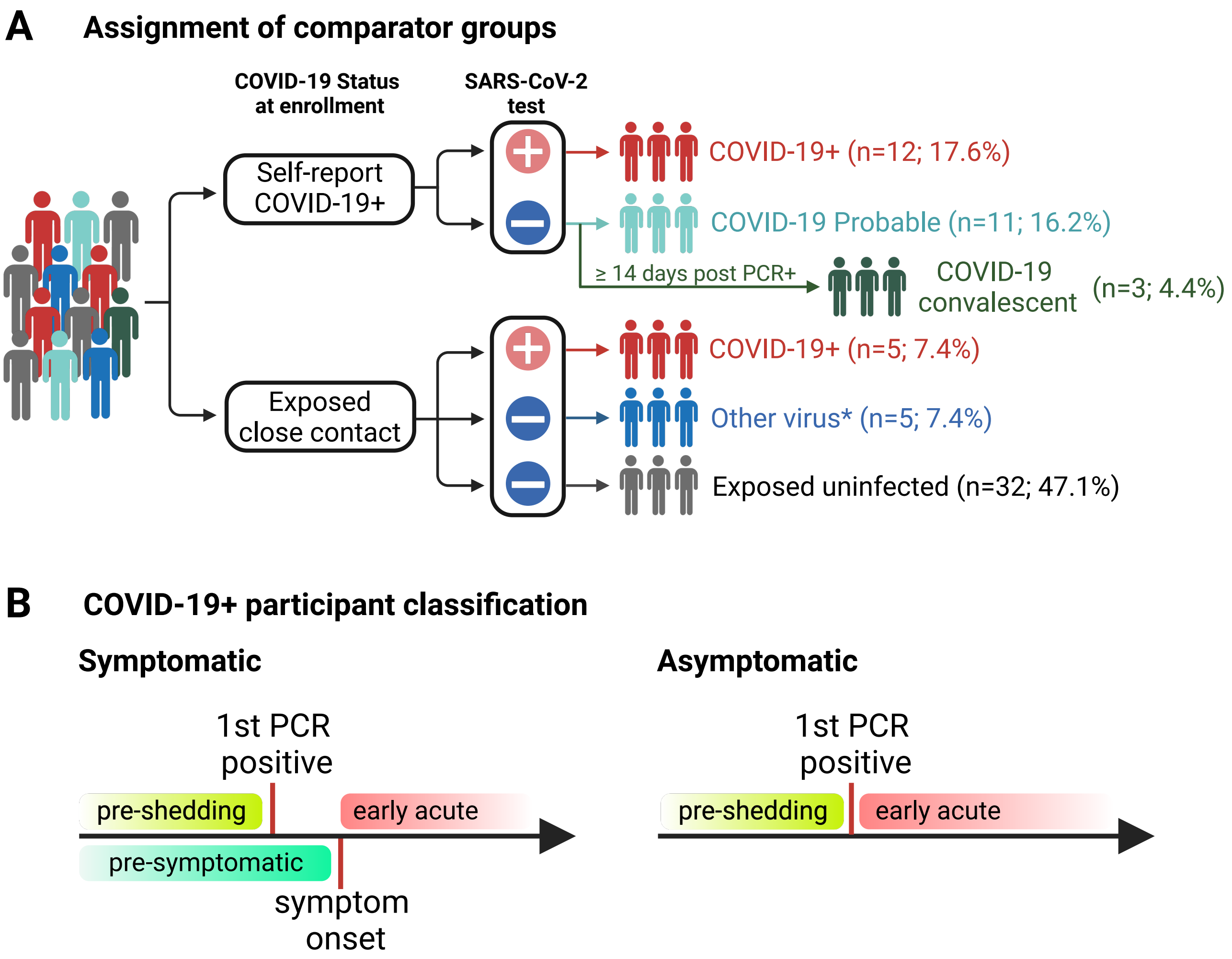


**Figure S1. Participant classification**. A) Comparator group assignment based on nasal swab test results B) Classification of symptomatic and asymptomatic COVID-19+ participants based on timing of their first sample collection day (colored boxes).

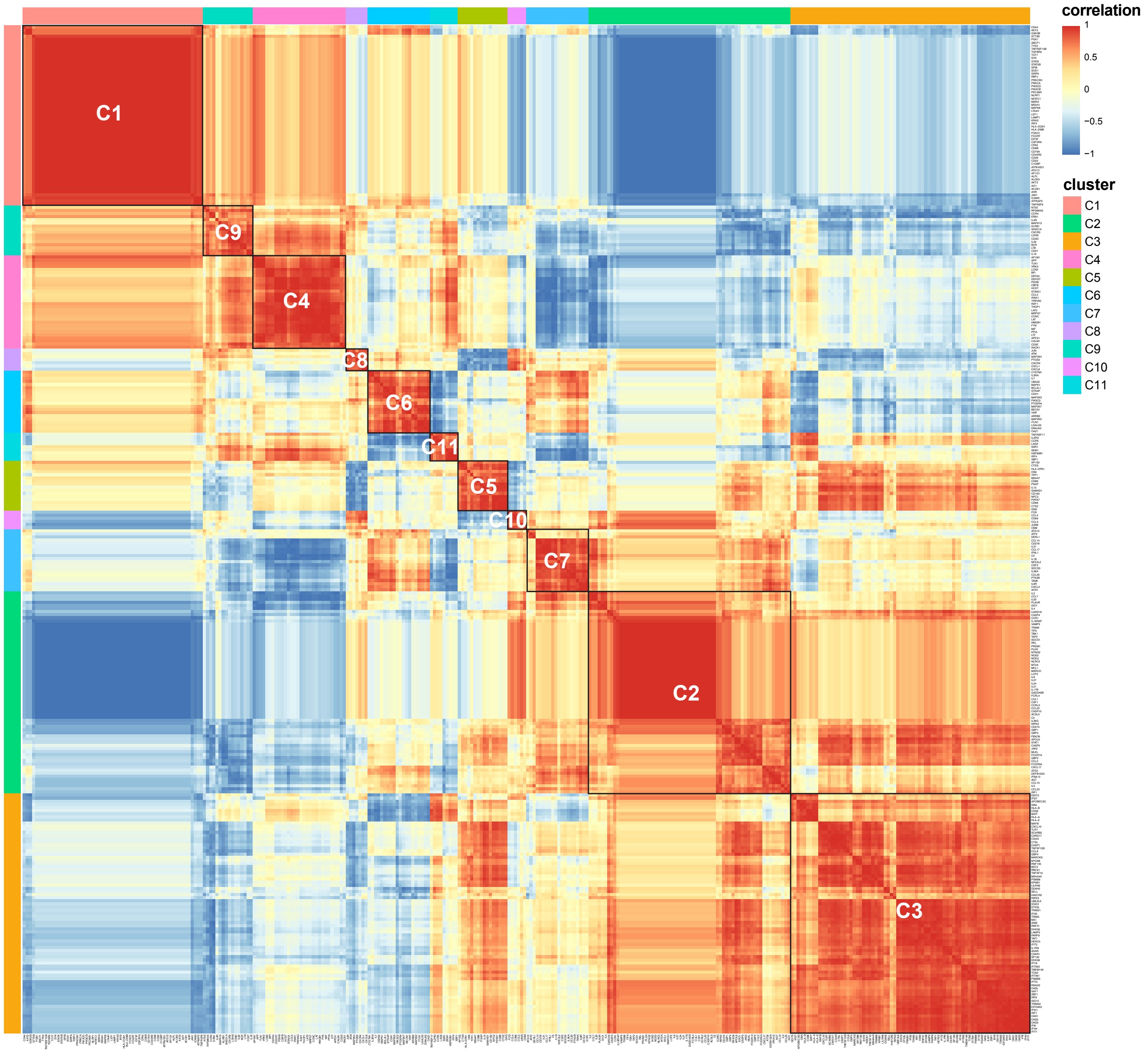


**Figure S2. Hierarchical clustering based on Spearman correlations between GAMM smooth fits of dynamic genes identified during SARS-CoV-2 infection.**

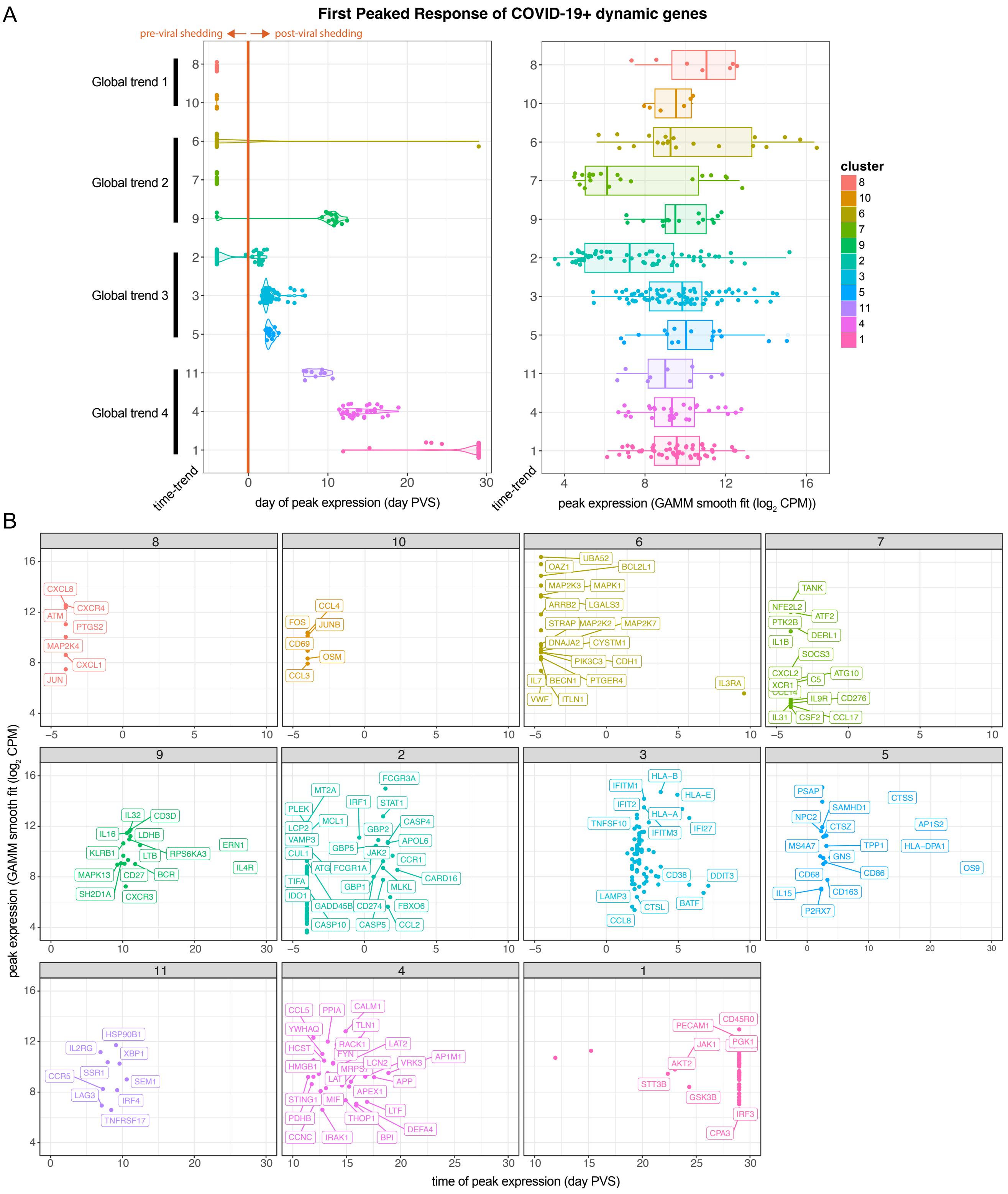


**Figure S3. First peaked response of significant COVID-19+ dynamic genes across the eleven longitudinal gene clusters identified through hierarchical clustering.** A) timing (violin plot) and level (box plot) of peak gene expression observed from -4 to 28 days PVS. B) Membership of COVID-19+ dynamic genes within the eleven time-trend clusters.

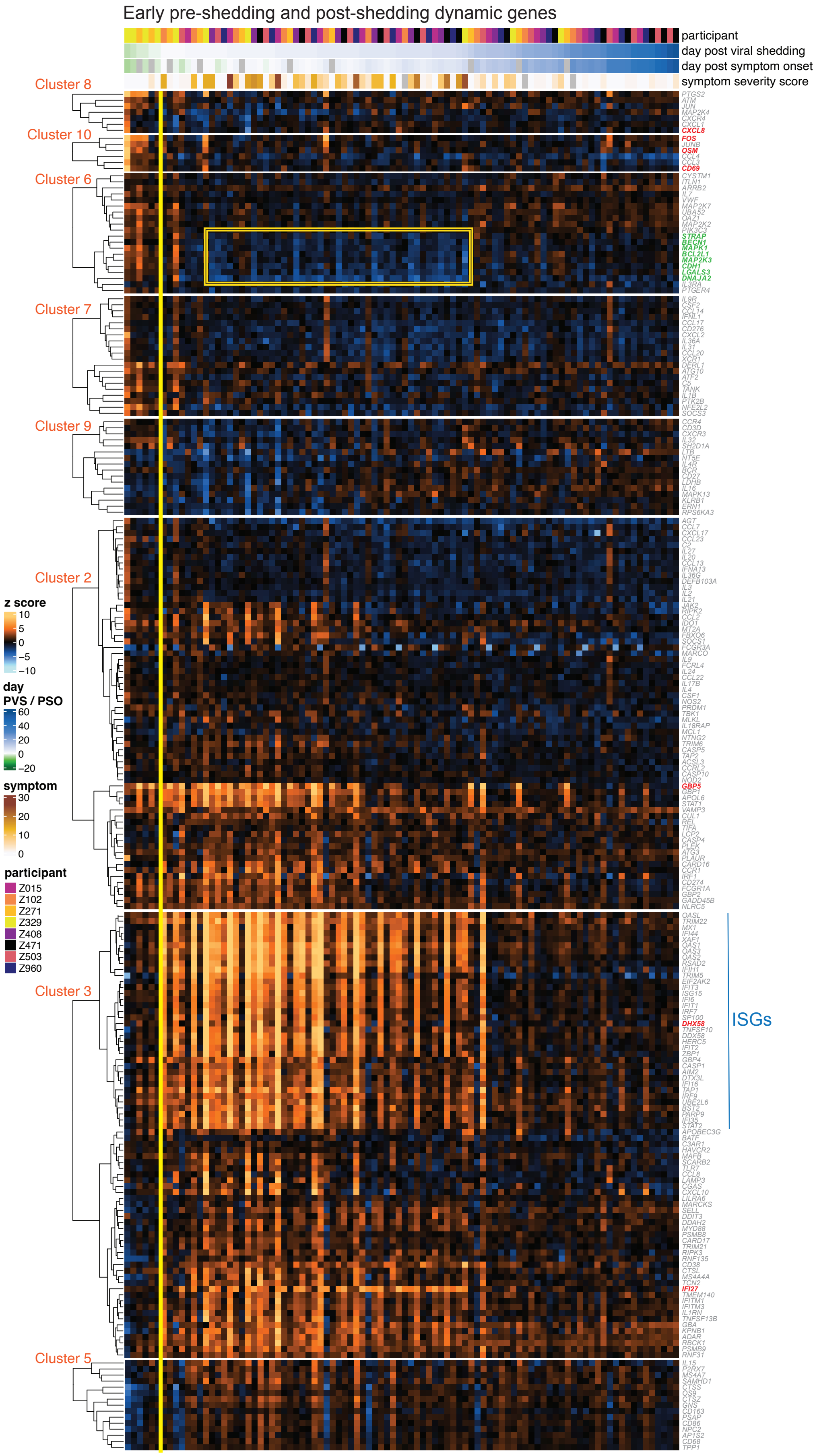
Early pre-shedding and post-shedding dynamic genes
participant
day post viral shedding
day post symptom onset
symptom severity score
Cluster 8
Cluster 10
Cluster 6
Cluster 7
Cluster 9
Cluster 2
Cluster 3
Cluster 5
z score
10
5
0
−5
−10
day
PVS / PSO
60
40
20
0
−20
symptom
30
20
10
0
participant
Z015
Z102
Z271
Z329
Z408
Z471
Z503
Z960
ISGs
CXCL8
FOS
OSM
CD69
STRAP
BECN1
MAPK1
BCL2L1
MAP2K3
CDH1
LGALS3
DNAJA2
GBP5
DHX58
IFI27

**Figure S4. Temporal kinetics of early pre- and post-shedding dynamic genes.** A) Heatmap depicting hierarchical clustering of early dynamic genes (rows) within each early time-trend cluster in COVID-19+ participants. Heatmap columns represent participant samples ordered by days post viral shedding (day PVS). Gene expression z scores (heatmap color) were calculated using the mean and standard deviations measured from unexposed-healthy controls ($n_{participants} = 8$; $n_{samples} = 39$). Yellow vertical line separates pre-shedding and post-shedding samples. For each participant sample, the collection day relative to the participant's first symptom onset date (days PSO) and first detectable viral shedding (days PVS), and the total daily symptom burden score for the same collection day are annotated above the heatmap.

Parent GO biological processes terms enriched

**early pre-shedding genes**

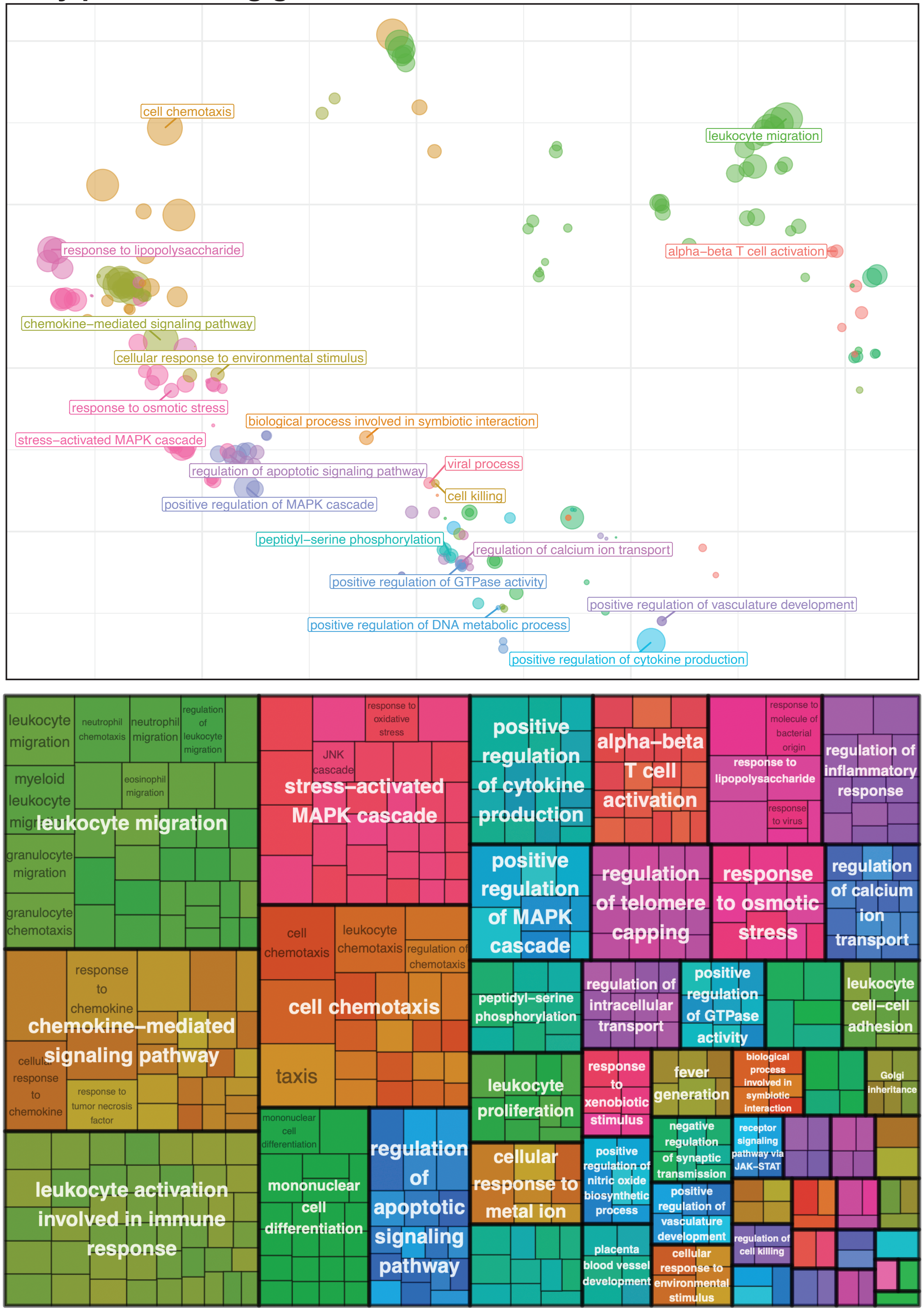

**Figure S5. Gene Ontology (GO) enrichment analysis of early pre-shedding dynamic genes.** Scatterplots (top) and treemaps (bottom) of reduced GO terms.

Parent GO biological processes terms enriched

**early post-shedding genes**

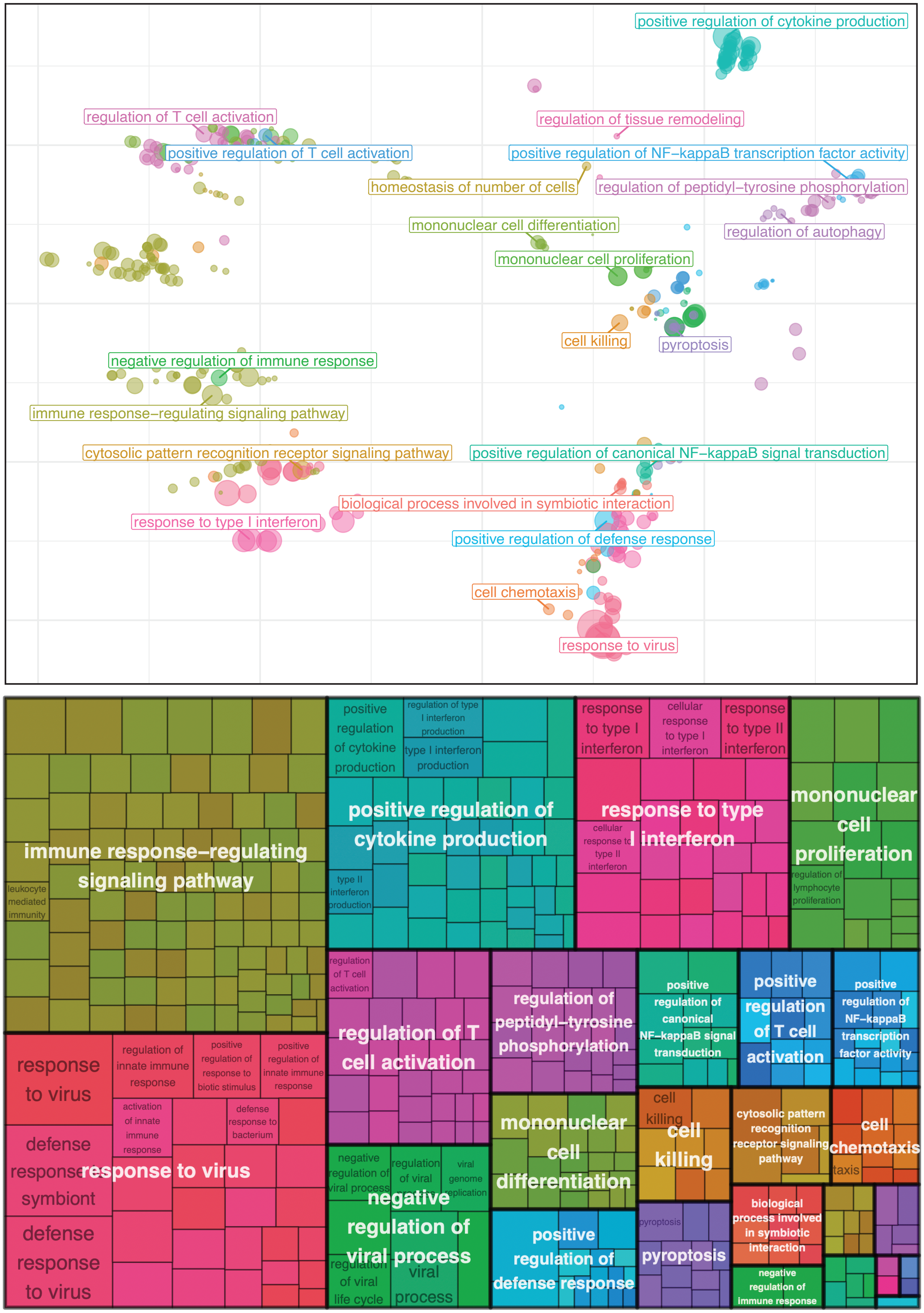

**Figure S6. Gene Ontology (GO) enrichment analysis of early post-shedding dynamic genes.** Scatterplots (top) and treemaps (bottom) of reduced GO terms.

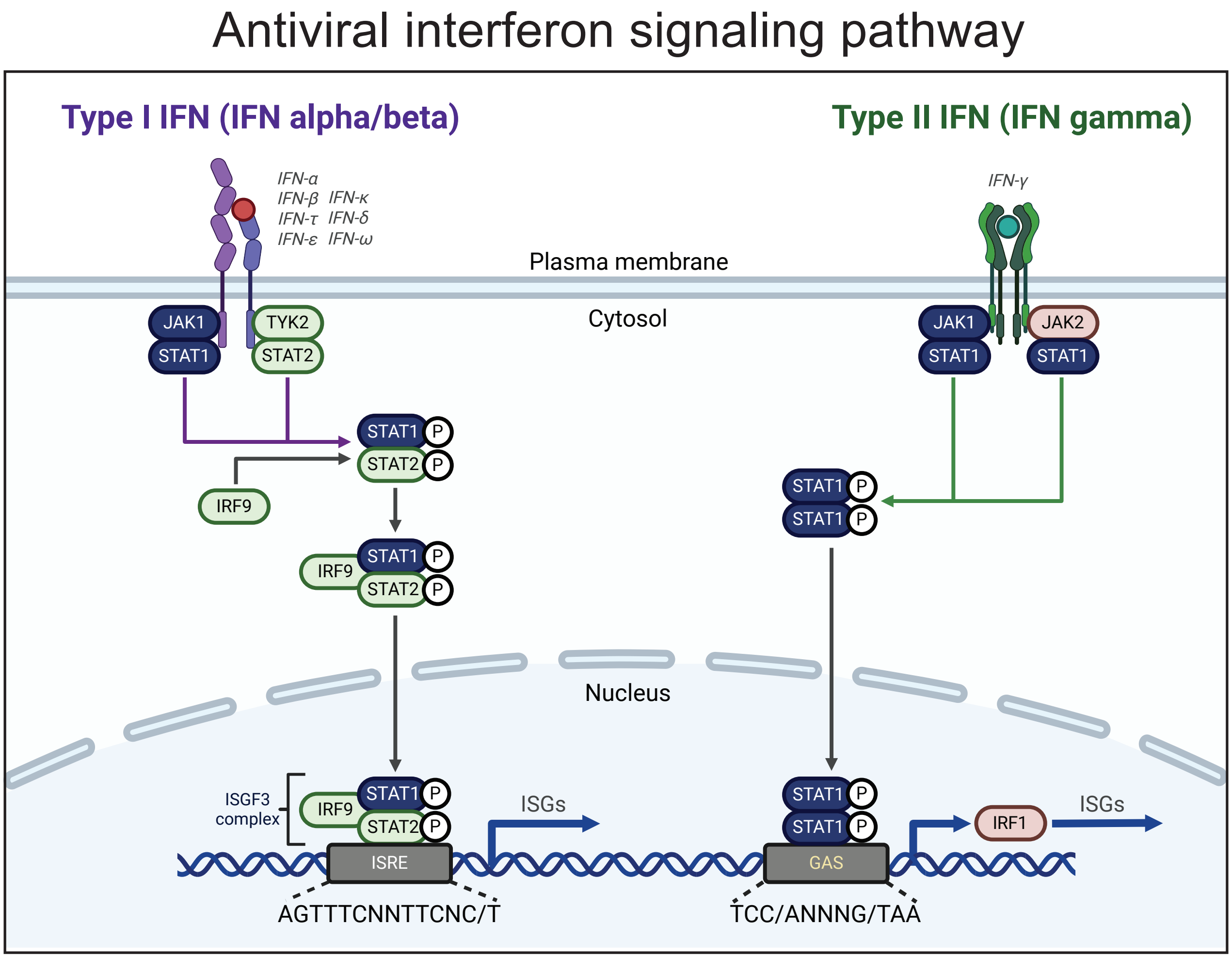


**Figure S7.** Interferon alpha/beta and gamma signaling pathway. Genes shared between both pathways were highlighted blue. Genes specific to IFNa/b signaling were highlighted in green while genes specific to IFNg signaling were highlighted in red.

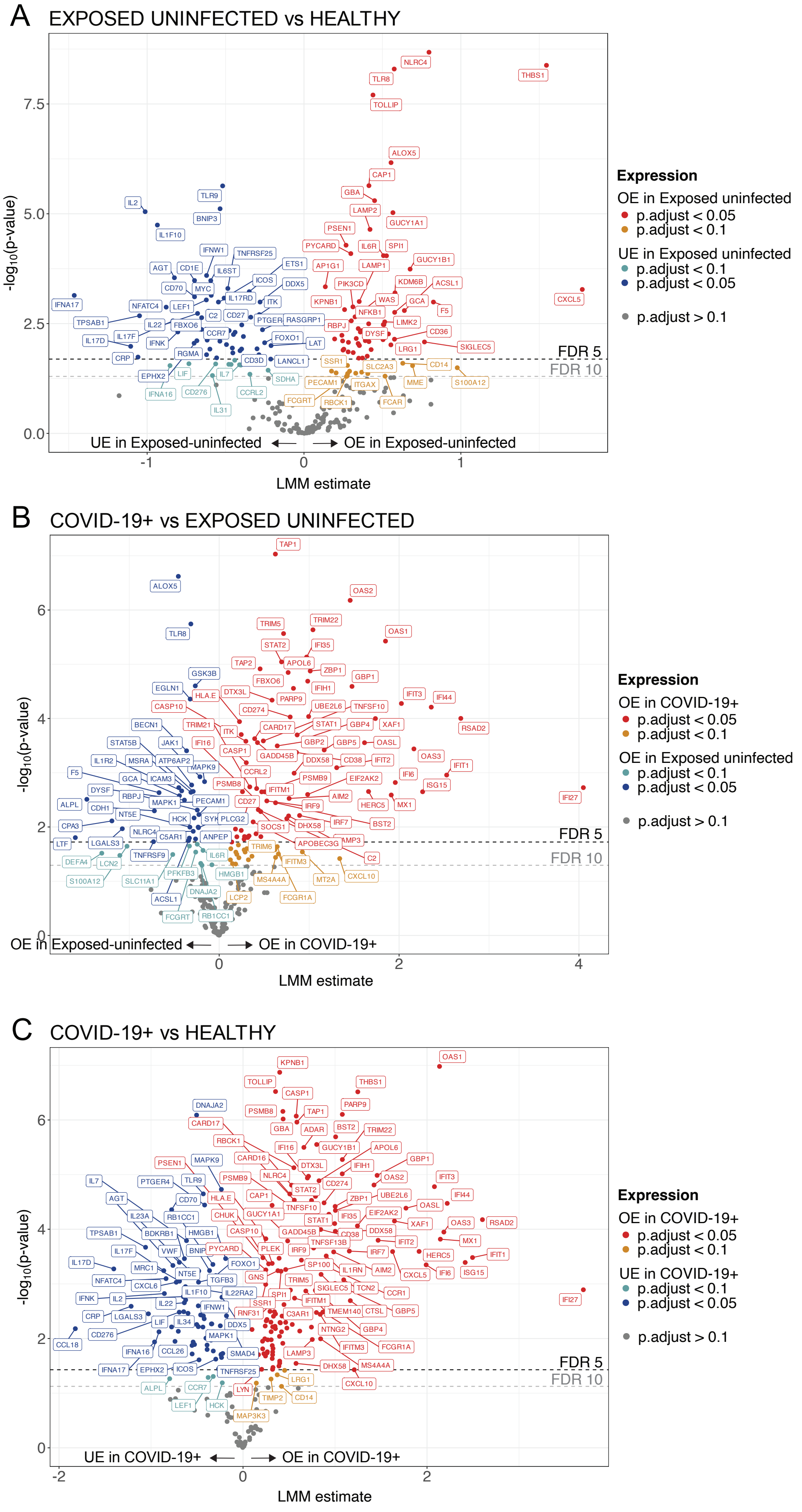

A
EXPOSED UNINFECTED vs HEALTHY
-log10(p-value)
LMM estimate
NLRC4
TLR8
THBS1
TOLLIP
ALOX5
CAP1
GBA
LAMP2
GUCY1A1
PSEN1
PYCARD
IL6R
SPI1
AP1G1
LAMP1
GUCY1B1
PIK3CD
KDM6B
ACSL1
KPNB1
WAS
GCA
CXCL5
F5
RBPJ
NFKB1
LIMK2
DYSF
CD36
LRG1
SIGLEC5
SSR1
SLC2A3
CD14
PECAM1
ITGAX
MME
S100A12
FCGRT
RBCK1
FCAR
TLR9
IL2
BNIP3
IL1F10
IFNW1
TNFRSF25
AGT
CD1E
IL6ST
ETS1
CD70
MYC
ICOS
DDX5
IL17RD
IFNA17
NFATC4
LEF1
ITK
C2
CD27
PTGER
RASGRP1
TPSAB1
IL22
FBXO6
CCR7
FOXO1
IL17D
IL17F
IFNK
LAT
RGMA
CRP
CD3D
LANCL1
EPHX2
LIF
IL7
SDHA
IFNA16
CD276
CCRL2
IL31
UE in Exposed-uninfected
OE in Exposed-uninfected
FDR 5
FDR 10
Expression
OE in Exposed uninfected
p.adjust < 0.05
p.adjust < 0.1
UE in Exposed uninfected
p.adjust < 0.1
p.adjust < 0.05
p.adjust > 0.1
B
COVID-19+ vs EXPOSED UNINFECTED
TAP1
ALOX5
OAS2
TLR8
TRIM5
TRIM22
OAS1
STAT2
IFI35
TAP2
APOL6
ZBP1
GSK3B
FBXO6
GBP1
EGLN1
HLA.E
DTX3L
PARP9
IFIH1
IFIT3
IFI44
CASP10
CD274
UBE2L6
TNFSF10
BECN1
TRIM21
CARD17
STAT1
GBP4
XAF1
RSAD2
ITK
STAT5B
JAK1
IFI16
GBP2
GBP5
OASL
CASP1
IL1R2
MSRA
ATP6AP2
GADD45B
OAS3
DDX58
CD38
IFIT2
IFIT1
F5
MAPK9
CCRL2
GCA
ICAM3
PSMB8
PSMB9
IFI6
EIF2AK2
ISG15
DYSF
IFITM1
AIM2
RBPJ
IFI27
ALPL
MAPK1
PECAM1
CD27
IRF9
HERC5
MX1
CDH1
NT5E
HCK
SYK
PLCG2
IRF7
BST2
CPA3
NLRC4
SOCS1
DHX58
C5AR1
ANPEP
LTF
LGALS3
APOBEC3G
LAMP3
TRIM6
TNFRSF9
IL6R
C2
DEFA4
LCN2
IFITM3
PFKFB3
HMGB1
S100A12
SLC11A1
MS4A4A
MT2A
CXCL10
DNAJA2
ACSL1
LCP2
FCGR1A
FCGRT
RB1CC1
OE in Exposed-uninfected
OE in COVID-19+
FDR 5
FDR 10
Expression
OE in COVID-19+
p.adjust < 0.05
p.adjust < 0.1
OE in Exposed uninfected
p.adjust < 0.1
p.adjust < 0.05
p.adjust > 0.1
C
COVID-19+ vs HEALTHY
KPNB1
OAS1
TOLLIP
THBS1
CASP1
DNAJA2
PSMB8
TAP1
PARP9
CARD17
GBA
ADAR
BST2
TRIM22
RBCK1
IFI16
GUCY1B1
APOL6
PSEN1
CARD16
GBP1
MAPK9
DTX3L
IFIH1
IL7
PTGER4
TLR9
PSMB9
NLRC4
OAS2
IFIT3
CD274
AGT
CD70
HLA.E
STAT2
CAP1
ZBP1
UBE2L6
IFI44
IL23A
RB1CC1
CHUK
TNFSF10
OASL
GUCY1A1
IFI35
EIF2AK2
STAT1
XAF1
OAS3
RSAD2
TPSAB1
BDKRB1
HMGB1
CASP10
GADD45B
DDX58
CD38
MX1
IL17F
VWF
BNIP
PYCARD
TNFSF13B
PLEK
IRF9
IFIT2
IRF7
HERC5
IFIT1
IL17D
FOXO1
SP100
MRC1
NT5E
IL1RN
AIM2
IFI6
CXCL5
ISG15
NFATC4
CXCL6
TGFB3
GNS
TRIM5
SIGLEC5
TCN2
CCR1
IL1F10
IL22RA2
SPI1
IFNK
IL2
IFITM1
CTSL
GBP5
IL22
SSR1
IFNW1
TMEM140
RNF31
C3AR1
CRP
LGALS3
LIF
IL34
DDX5
NTNG2
GBP4
MAPK1
CD276
CCL18
IFITM3
FCGR1A
IFNA16
CCL26
SMAD4
LAMP3
IFNA17
EPHX2
ICOS
TNFRSF25
DHX58
MS4A4A
CXCL10
ALPL
CCR7
LRG1
LYN
LEF1
HCK
TIMP2
CD14
MAP3K3
UE in COVID-19+
OE in COVID-19+
FDR 5
FDR 10
Expression
OE in COVID-19+
p.adjust < 0.05
p.adjust < 0.1
UE in COVID-19+
p.adjust < 0.1
p.adjust < 0.05
p.adjust > 0.1

**Figure S8. Exposed-uninfected individuals portrayed robust differential response**. Volcano plot depicting overexpressed (estimate > 0) and underexpressed (estimate < 0) genes in A) "exposed-uninfected vs healthy", B) "COVID-19+ vs exposed-uninfected", and "COVID-19+ vs healthy" participants. x-axis shows the LMM estimates between each pairwise contrast. y-axis shows the $-log_{10}$(p-value) derived from the comparison. Horizontal dotted lines denote genes with adjusted p-values < 0.05 (FDR 5) and adjusted p-values < 0.1 (FDR 10).

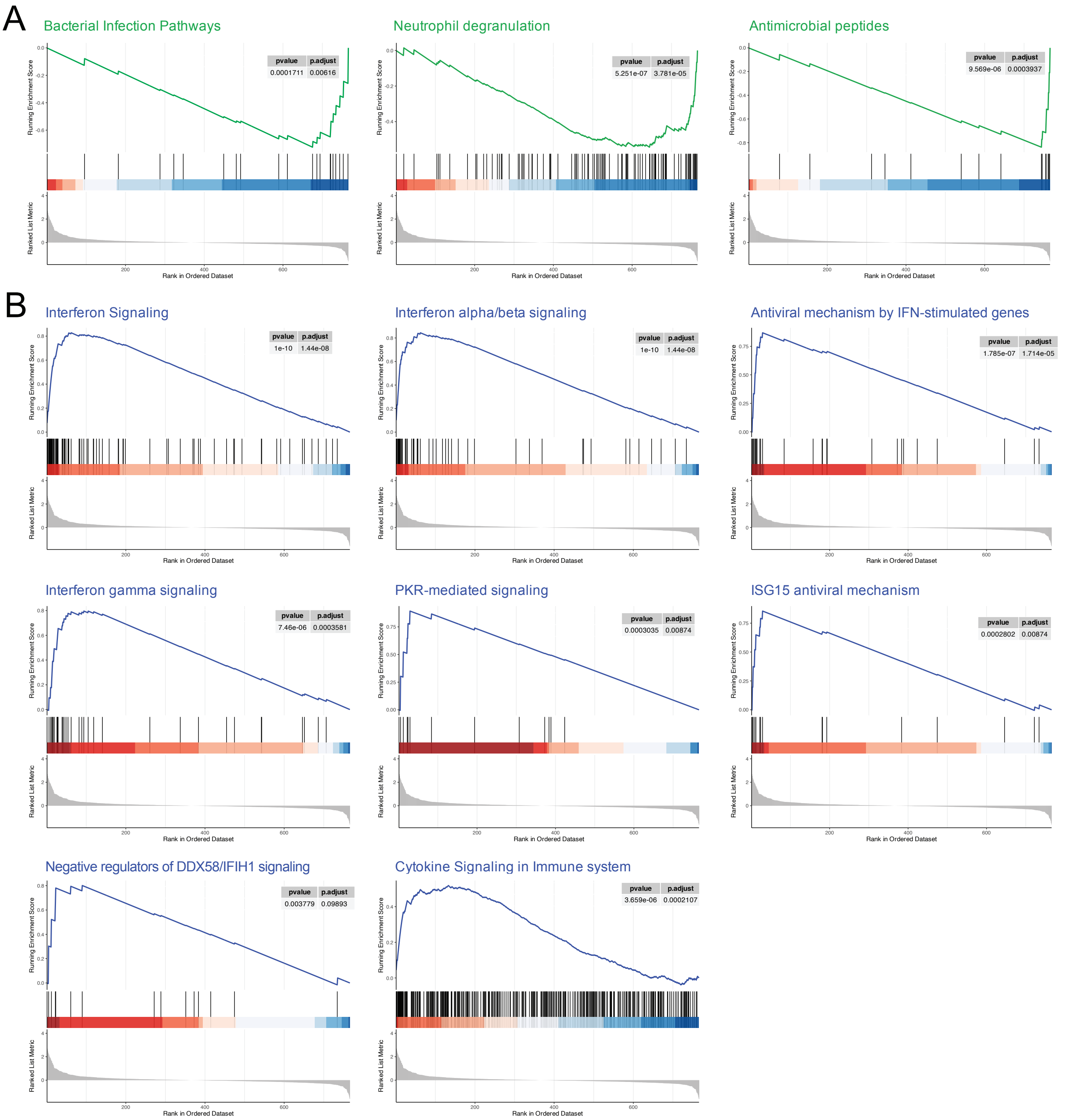


**Figure S9. Exposed-uninfected individuals showed increased expression of antimicrobial peptides.** A) GSEA plot showing overexpression of neutrophil degranulation, bacterial infection, and antimicrobial peptide pathway genes in exposed-uninfected participants. B) GSEA plot showing overexpression of antiviral mechanisms in COVID-19+ participants.

### SUPPLEMENTAL TABLES

**Table S1. Reasons for ineligibility**

| Reason for screen fail | | % |
|---|---|---|
| Under 18 years of age/no age reported | 13 | 1.2 |
| Immunosuppressed, pregnant, prisoner | 102 | 9.1 |
| COVID-19+ between 2 weeks - 6 months | 190 | 17 |
| previously vaccinated/previously vaccinated and no household exposure[1] | 632 | 56 |
| no exposure | 41 | 3.6 |
| currently symptomatic | 99 | 8.8 |
| symptom resolved | 18 | 1.6 |
| Incomplete screen survey response | 29 | 2.6 |
| Total number of screen fails | 1124 | |

[1] Exposed individuals with unknown infection status at the time of screening were excluded if they had been fully vaccinated to enrich for positive cases (infected participants). Vaccinated individuals with known COVID-19 infection at the time of screening were not excluded. In February 2022, exposed individuals with unknown infection status at the time of screening were excluded if they have been fully vaccinated and were exposed to a non-household member. This protocol modification was adapted to accommodate the increased number breakthrough infections observed during the later enrollment period.

**Table S2. List of respiratory pathogens screened in the OpenArray analysis platform**

| **Pathogen Category** | **Strains** |
|---|---|
| Influenza | Pan Flu A, Flu A (H3N2), Flu A (H1N1), Pan Flu B, Pan Flu C |
| Parainfluenza | Human Parainfluenza 1 - 4 |
| Enterovirus | Pan enterovirus and Enterovirus D68 |
| Rhinovirus | Rhinovirus 1 and 2 |
| Adenovirus | Adenovirus 1 and 2 |
| non-SARS coronavirus | HKU1, NL63, 229E, OC43 |
| SARS coronavirus | SARS-CoV-2 |
| Respiratory syncytial virus | RSV A and RSV B |
| Metapneumovirus | hMPV |
| Bacterial pneumonia | *Streptococcus pneumoniae, Mycoplasma pneumoniae, Chlamydia pneumoniae* |
